# Rapid Molecular Assessment of Kidney Allograft Rejection Using the Biopsy Transport Medium

**DOI:** 10.64898/2026.10.01.26364480

**Authors:** Ziyang Li, Nienke Kalverboer, Danny van der Helm, Giulia Callegaro, Bob van de Water, Soufian Meziyerh, Aiko P.J. de Vries, Hans J. Baelde, Jesper Kers

## Abstract

**Introduction:** Timely diagnosis of kidney allograft rejection is critical for post-transplant management, yet treatment decisions may need to be made empirically while awaiting the final pathology report. We previously demonstrated that biopsy transport medium (BTM) contains molecular signals reflective of allograft rejection. Building on these findings, we developed a rapid BTM–qPCR workflow using a targeted gene panel for early molecular assessment of rejection.

**Methods:** Kidney transplant biopsies assessed at Leiden University Medical Center from April 2023 to November 2024 were eligible. A predefined gene panel was quantified in BTM by qPCR, while matched formalin-fixed paraffin-embedded tissues were profiled using Tempo-Seq. Elastic Net, Random Forest, and support vector machine models were developed to classify rejection and predict continuous histological indices directly from the BTM.

**Results:** A total of 134 paired BTM samples and biopsy tissues were analyzed. The best-performing BTM gene-panel models demonstrated good discrimination for rejection, with a median AUC of 0.80, and good calibration (median slope, 0.92; median intercept, 0.08). For continuous histological indices, the best-performing BTM gene-panel models achieved R² values of 0.33 for Activity, 0.31 for TCMR/TI, and 0.27 for AMR/MVI. Corresponding biopsy gene-panel models showed similar patterns across diagnostic outcomes.

**Conclusions:** The BTM–qPCR workflow captures molecular features of kidney allograft rejection and enables prediction of rejection status along the spectrum of continuous activity indices. By providing same-day molecular information while definitive biopsy interpretation is pending, this rapid approach has the potential to complement conventional histopathological diagnostics and support early clinical decision-making.

**Lay Summary:** Kidney transplantation is the optimal treatment for end-stage kidney disease, and effective post-transplant management is critical for long-term survival of the transplanted kidney. Therefore, prompt treatment decisions and timely interventions are essential. However, a complete and final pathological assessment of a kidney biopsy can sometimes take several days. A molecular diagnostic approach using qPCR on biopsy transport medium (BTM), the liquid used to transport biopsy samples, has the potential to complement conventional pathological assessment by providing rapid information to support clinical decision-making. Our study included 134 kidney transplant biopsy samples and measured the expression of a panel of 19 genes in BTM. These gene expression measurements were able to distinguish rejection from non-rejection and predict continuous histological indices. Our findings suggest that, while pathological assessment is pending, the BTM–qPCR approach could provide same-day molecular information to support earlier clinical decision-making.

## Introduction

Kidney allograft rejection remains a primary concern following transplantation, as it is strongly associated with poor long-term outcomes^1–3^. Timely identification of post-transplant kidney allograft status is critical for informing clinical decision-making and guiding prompt intervention. The gold standard for diagnosis is the histopathological assessment of biopsy samples^4,5^. This process relies on expert interpretation by nephropathologists; however, it is often hindered by significant interobserver variability across clinical centers^6,7^. This has become one of the major reasons why emerging molecular diagnostics and digital pathology have gained traction and show promise in complementing traditional histology^5,8–10^. Nevertheless, these methods often lack the rapid turnaround time necessary for clinical utility as they typically require at least overnight fixation. This creates a significant “diagnostic gap”, for instance, a biopsy performed on a Friday may not yield a definitive diagnosis until the following Monday. During this interval, clinicians are often forced to rely on empirical, experience-based treatment, which can lead to suboptimal outcomes, irreversible graft damage, or even graft loss^11,12^

To address these challenges, we recently proposed a novel liquid biopsy approach utilizing Biopsy Transport Medium (BTM)^13^. Historically, the BTM is a discarded byproduct of the transport process, serving only to preserve the biopsy tissue during its transit from the site of biopsy to the pathology laboratory. We found that cells released from the tissue into the BTM during transport contain valuable biological information. In a proof-of-concept study involving 12 transplant samples, we successfully extracted RNA from BTM samples and demonstrated its suitability for gene expression profiling using transcriptomics (whole transcriptome RNA-sequencing and the NanoString Banff Human Organ Transplant targeted panel) and quantitative real-time PCR (qPCR). We identified differentially expressed genes associated with rejection status and specific rejection phenotypes.

Recognizing that transcriptomic gene expression profiling cannot provide same-day turnaround, the current study aims to validate a targeted gene panel using qPCR to facilitate rapid, diagnostic assessment of kidney allograft status. This approach offers several distinct advantages over existing diagnostic methods. First, qPCR is a rapid, standardized, and cost-effective procedure that enables same-day preliminary results. Second, compared to liquid biopsies such as blood or urine, which are often confounded by systemic biological noise, the BTM more directly captures local molecular and immune processes within the allograft.

Third, while existing molecular diagnostic techniques requiring additional biopsy cores or FFPE sections and are both costly and time-consuming, our method provides a scalable, low-cost, and time-efficient alternative, addressing a critical barrier to widespread clinical translation, including adoption in lower resource settings.

## Methods

### Study design and patient cohort

All kidney transplant biopsy assessed at Leiden University Medical Center from April 2023 to November 2024 were eligible for inclusion. Clinical data were retrieved from LUMC Biobank Kidney Transplantation (NTX). A dedicated data manager is in charge of extracting the relevant data for this study. This study was approved by the LUMC Review Committee Biobank & Biomaterials (TCBio) under number RP24.068.

### Clinical outcome definition

The primary outcome was a binary diagnosis of rejection versus non-rejection. Rejection was defined according to the 2024 Banff classification based on histopathological evaluation of kidney transplant biopsies^14^. Samples obtained at the time of biopsy-proven rejection were classified as rejection samples, whereas samples without histological evidence of rejection, but other abnormal changes were classified as non-rejection. The secondary outcome comprised continuous indices derived from Banff lesion scores, which characterize the phenotypic spectrum of kidney transplant rejection. These included the AMR/MVI Index, TCMR/TI Index, Activity Index, and Chronicity Index^15^.

### Sample collection and processing

BTM samples were collected from kidney transplant biopsies submitted for routine diagnostic evaluation. The samples were centrifuged at 3000g to obtain the cell pellet, which was subsequently lysed with 350 μl buffered RLT (included in RNeasy Micro Kit, QIAGEN, Hilden, Germany) supplemented with 3.5 μl 2-Mercaptoethanol (Merck Schuchardt OHG, Hohenbrunn, Germany). Total RNA was subsequently isolated using the RNeasy Micro Kit according to the manufacturer’s instructions. The RNA was reverse transcribed into cDNA using AMV reverse transcriptase (Roche, Basel, Switzerland) and random hexamer primers. qPCR analysis was performed using IQ^TM^SYBR Green Supermix (Bio-Rad, Hercules, CA, USA) in conjunction with the Bio-Rad CFX96 Touch Real-Time PCR Detection System and the Bio-Rad CFX Maestro 2.2 (5.2.008.0222) software.

The corresponding FFPE tissue were retrieved from the archives of the Department of Pathology. Whole-transcriptome gene expression profiling was performed using the TempO-Seq® FFPE Whole Transcriptome Assay (BioSpyder Technologies, Carlsbad, CA, USA) according to the manufacturer’s instructions^16^. Raw sequencing reads were processed using the TempO-Seq analysis pipeline to generate gene-level count data for downstream normalization and statistical analyses.

### Gene expression measurements and data preprocessing

#### Predefined gene panel

The targeted gene panel was predefined based on our previous study and supporting evidence from the literature. Genes were selected according the following criteria: 1) differential expression between rejection and non-rejection, as well as between rejection phenotype (AMR and TCMR); 2) present in priority gene list reported in the Banff 2017 Kidney Meeting Report^17^. In addition, the panel was designed to capture the *cellular composition* and inflammatory state of the kidney allograft by including markers of major immune and renal parenchymal cell types, as well as genes associated with inflammation and cellular activation. The selected gene panel is presented in Supplementary Table 1.

#### qPCR analysis of BTM samples

Four housekeeping genes from B-HOT panel were selected and the expression stability was evaluated using the “Reference Gene Selection Tool” which is integrated in the Bio-Rad CFX Maestro 2.2 (5.2.008.0222) software. Raw qPCR data were analyzed using cycle threshold (Ct) values. The average Ct of the housekeeping genes was used as the reference for normalization. For each target gene, ΔCt was calculated as Ct_target − mean Ct_housekeeping, and relative expression was quantified as 2^−ΔCt^.

#### TempoSeq targeted whole transcriptomic profiling of the corresponding FFPE tissue

All probes with < 2 counts per million (CPM) in >98% of the samples are removed. Batch effects were corrected using the ComBat algorithm. Normalized expression values were subsequently calculated as CPM using the edgeR package. To stabilize variance and reduce the influence of highly expressed genes, CPM values were log2-transformed after the addition of a pseudocount of 1 [log2(CPM + 1)]. The resulting log2CPM values were used for statistical analyses and predictive model development.

### Predictive modeling framework

Prediction models were developed using three datasets: BTM gene-panel models, based on qPCR measurements of the predefined gene panel in BTM samples; biopsy gene-panel models, based on the corresponding genes measured by TempO-Seq in biopsy samples; and biopsy whole-transcript models, based on all transcripts measured by TempO-Seq in biopsy samples. The biopsy-derived models served as molecular references to assess the predictive capacity of the predefined gene panel in biopsy tissue and the potential added value of broader transcriptomic profiling. Models were developed using Elastic Net (EN), Random Forest (RF), and support vector machines with a radial basis function kernel (SVM-RBF). Model training and internal validation were performed using repeated five-fold cross-validation with 20 repetitions, using a common resampling framework across algorithms.

Hyperparameters were optimized by grid search, with predictors centered and scaled where appropriate. For whole-transcriptome models, near-zero-and zero-variance features were removed and recursive feature elimination was performed within the training data to prevent information leakage. Full details of preprocessing, hyperparameter grids, and feature-selection procedures are provided in the Supplementary Methods.

Performance was evaluated using out-of-fold predictions from the optimally tuned models. For binary diagnosis, discrimination was assessed using ROC AUC, accuracy, precision, recall, and F1-score; calibration was evaluated using calibration plots, calibration intercept and slope, and the Brier score. Clinical utility was assessed using decision curve analysis (DCA) across clinically relevant probability thresholds, with net benefit compared with treat-all and treat-none strategies. For continuous outcomes, performance was evaluated the coefficient of determination (R²), root mean square error (RMSE), mean absolute error (MAE), with Pearson (r) and Spearman (ρ) correlation coefficients additionally used to assess the association between predicted and observed values.

For model interpretation, gene-level coefficients or variable importance scores were extracted from the final optimally tuned models. EN coefficients were additionally expressed as odds ratios, whereas scaled variable importance scores were obtained for RF and SVM-RBF models.

### Statistical analysis

Continuous variables were summarized as mean ± standard deviation (SD) or median with interquartile range (IQR), while categorical variables were summarized as frequencies and percentages. Comparisons between groups were performed using Wilcoxon rank-sum test for continuous variables. Gene–gene correlations were assessed using Pearson correlation, whereas associations between gene expression and histopathological lesion scores were assessed using Spearman rank correlation. To assess whether the relative distances among genes and the patterns of gene–lesion associations were similar between BTM and biopsy, gene–gene correlation-derived distance matrices were compared using the Mantel test, while gene–lesion correlation patterns were compared using Pearson correlation. Statistical significance was assessed using two-sided P values, with P < 0.05 considered statistically significant. All statistical analyses were performed using R software (version 4.2.1; R Foundation for Statistical Computing, Vienna, Austria). Machine learning models were developed using the caret, glmnet, randomForest, and kernlab packages. Model performance was evaluated using the pROC and rmda packages, and data visualization was performed using the tidyverse and ggplot2 packages.

## Results

### Study population

A total of 133 kidney transplant recipients, corresponding to 134 kidney transplant biopsies and 134 paired BTM samples, were included in this study. The mean recipient age was 51 ± 14 years, and 58% of recipients were male. 43% received kidneys from deceased donors. 12% of recipients had undergone a previous kidney transplantation. The median cold ischemia time was 241[178, 633] minutes (Table 1).

**Table 1.** Baseline patient characteristics in the study population.

|  | N | Value |
| --- | --- | --- |
| **Recipient characteristics** |  |  |
| Age (y), mean (SD) | 133 | 51 (14) |
| Sex male, n (%) | 133 | 77 (58%) |
| **Cause of end stage renal disease, n (%)** | 134 |  |
| Autoimmune/vasculitic/vascular disease |  | 8 (6.0%) |
| Cystic/hereditary kidney disease |  | 26 (19%) |
| Diabetic/hypertensive/vascular nephropathy |  | 35 (26%) |
| Glomerulonephritis |  | 26 (19%) |
| Reflux/urologic nephropathy |  | 5 (3.7%) |
| Unknown etiology |  | 34 (25%) |
| **Donor characteristics** |  |  |
| Age (y), mean (SD) | 130 | 53 (16) |
| Sex male, n (%) | 128 | 58 (45%) |
| Deceased donor, n (%) | 133 | 56 (43%) |
| Expanded criteria donor, n (%) | 130 | 56 (43%) |
| **Transplant baseline characteristics** |  |  |
| Prior kidney transplant, n (%) | 133 | 16 (12%) |
| Cold ischemia time (minutes), mean (SD) | 128 | 214 (178, 633) |
| HLA-A/B/DR mismatch, mean (SD) number | 132 | 3 (3, 5) |
| DSA at the time of transplant, n (%) | 134 | 7 (5.2%) |

\* Two BTM samples miss clinical data.

A total of 98 biopsies (73%) were diagnosed with rejection, comprising 19 cases of AMR, 32 cases of TCMR, and 47 cases of mixed rejection. Among the 36 biopsies without rejection, the most frequent histological diagnoses included acute tubular injury/necrosis or tubulointerstitial nephritis (n = 27), interstitial fibrosis and tubular atrophy not otherwise specified ( n = 10), recurrent glomerulonephritis (n = 5), glomerulopathy (n = 1), calcineurin inhibitor (CNI) toxicity (n = 6), and BK polyomavirus nephropathy (n = 1). Most biopsies had more than one pathological diagnosis, a common phenomenon in kidney transplant biopsies. The distribution of biopsy diagnoses is presented in Figure 1.

**Figure 1.**
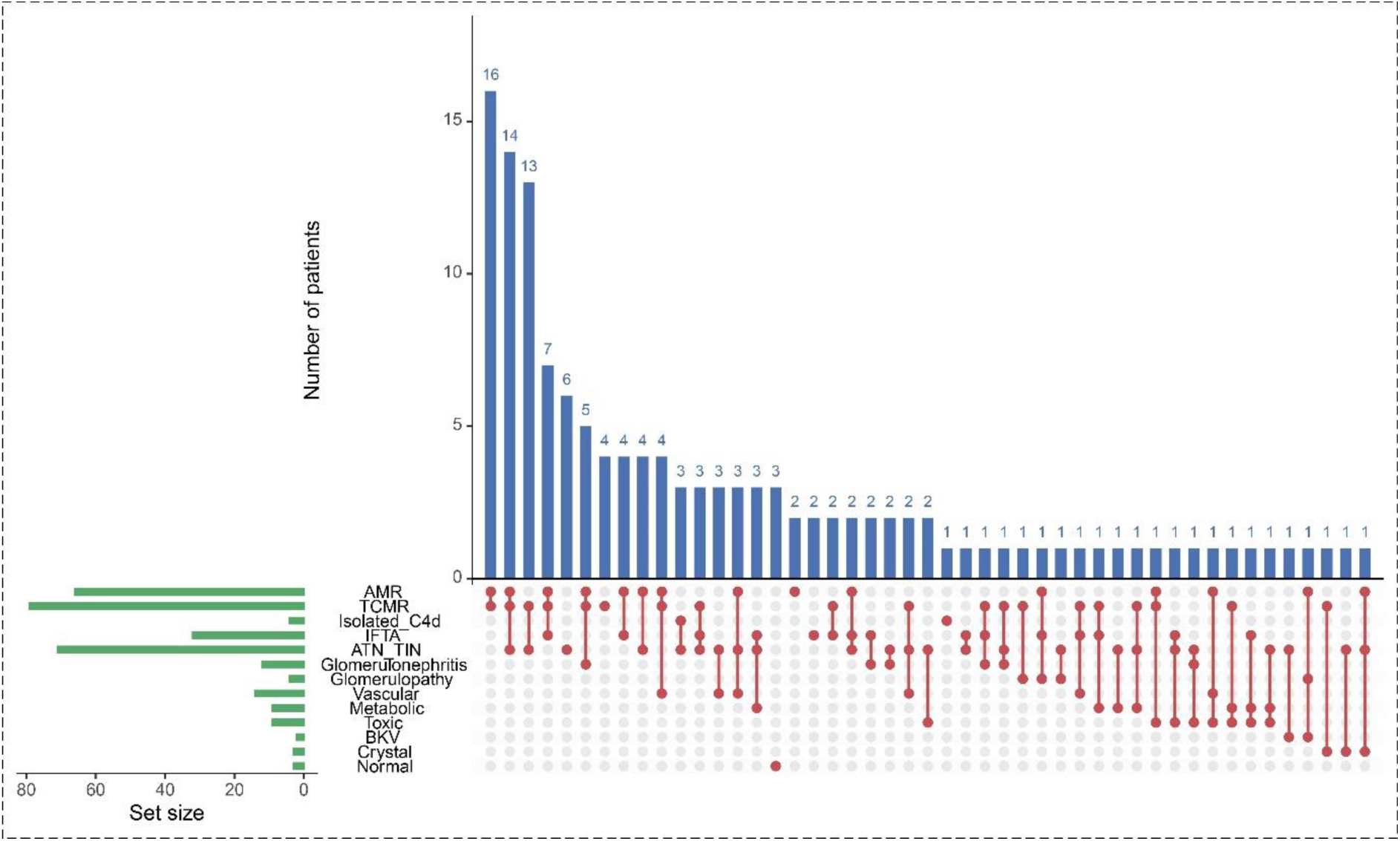
Distribution of diagnoses. The UpSet plot shows the distribution and overlap of diagnoses in the study cohort. Each dot represents a single diagnosis, and connected dots indicate diagnoses that co-occurred in the same biopsy. The upper bar plot shows the number of patients for each diagnosis combination, and the left bar plot shows the frequency of each individual diagnosis.

The histopathological characteristics of the biopsy cohort are summarized in Supplementary Table 2. The median (interquartile range) values of the AMR/MVI, TCMR/TI, Activity, and Chronicity indices were 2.72 (0.00, 5.35), 1.98 (0.39, 3.58), 5 (2, 8), and 3 (1, 6), respectively (Table 2). The distributions of individual Banff lesion scores and gene expression profiles derived from biopsy tissue and paired BTM samples are presented in Supplementary Figure 1.

**Table 2.**
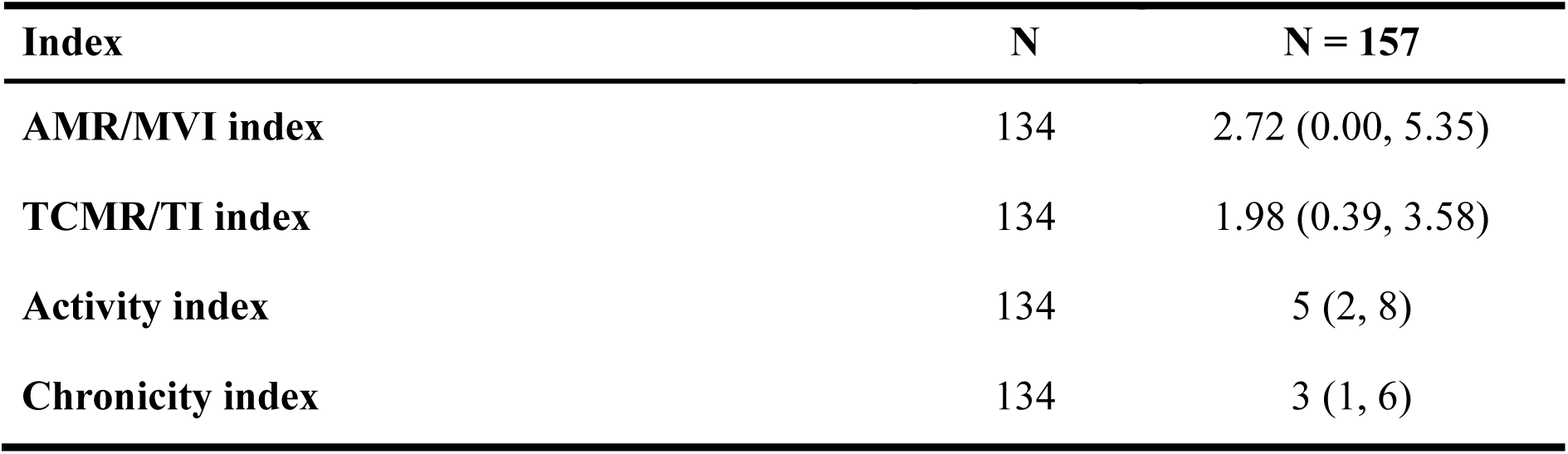
Continuous index characteristics in the study population.

### Association of predefined gene expression with Banff lesion scores

Spearman correlation analysis was performed to evaluate associations between gene expression and individual Banff lesion scores in paired BTM and biopsy samples (Figure 2a,b). In BTM, T-cell–associated markers (*CD3E* and *CD8A*) and *CTLA4* were predominantly associated with tubulointerstitial inflammatory lesions (i, t, and ti), whereas interferon-related genes *CXCL10* and *GBP1* showed stronger correlations with microvascular inflammation, particularly g and ptc. *CXCL10* showed the strongest correlations with g (ρ = 0.55) and ptc (ρ = 0.59), followed by *GBP1* (g: ρ = 0.45; ptc: ρ = 0.50), all *P* < 0.001. Biopsy-derived gene expression showed generally stronger correlations with tubulointerstitial inflammation, with *CD3E* showing the strongest associations with ti (ρ = 0.76), i (ρ = 0.69), and t (ρ = 0.68), all *P* < 0.001, followed by *CTLA4* and *CD8A*. Notably, *CXCL10* and *GBP1* were also strongly correlated with tubulointerstitial lesions (ti: ρ = 0.40 and 0.48; i: ρ = 0.47 and 0.49; t: ρ = 0.46 and 0.55, respectively; all *P* < 0.001), while retaining associations with microvascular inflammation, particularly ptc (*CXCL10*: ρ = 0.49; *GBP1*: ρ = 0.51; both *P* < 0.001).

**Figure 2.**
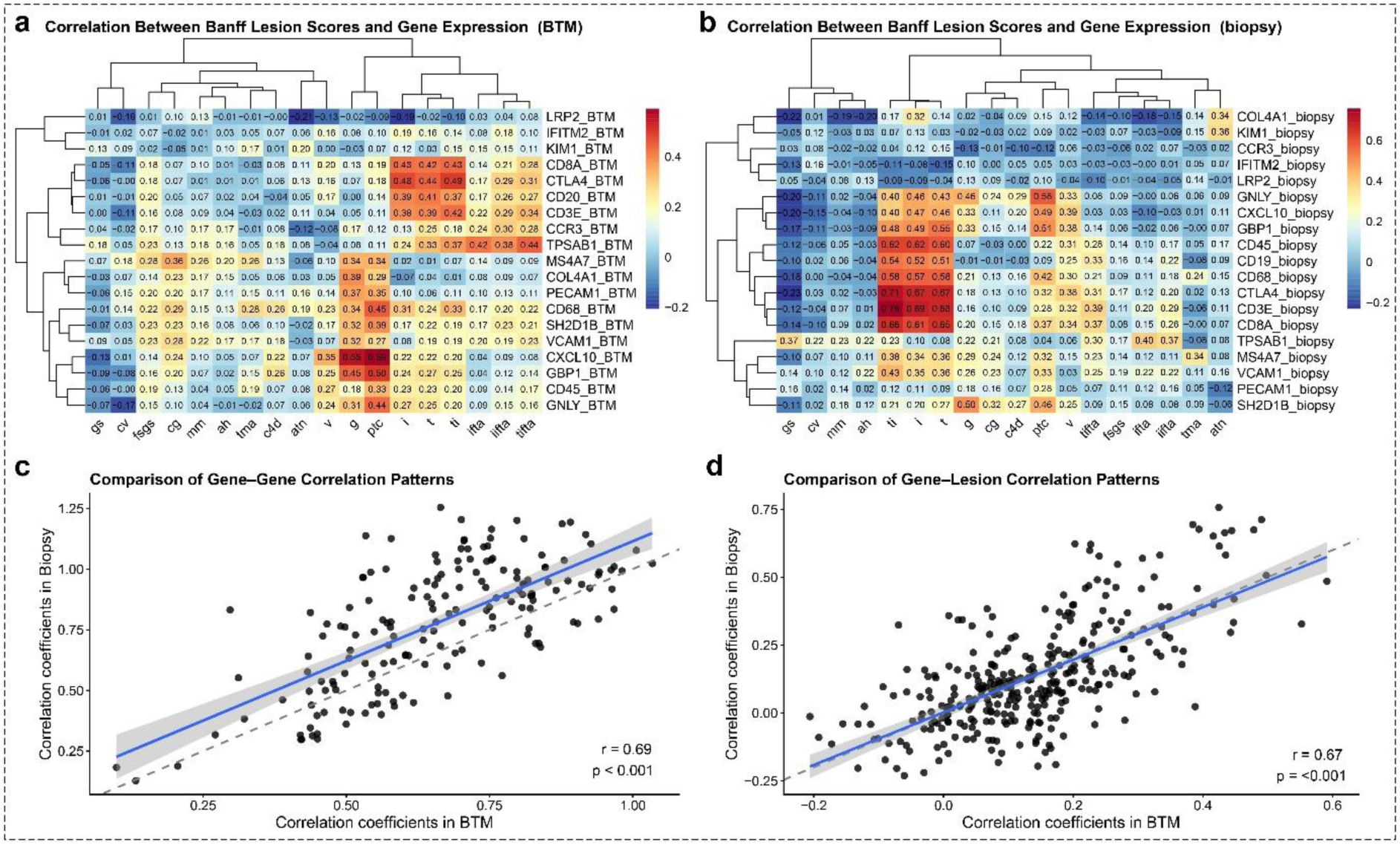
Concordance of gene–lesion and gene–gene correlation patterns between BTM and biopsy. a,b, Heatmaps showing Spearman correlation coefficients between Banff lesion scores and gene expression derived from BTM (**a**) and biopsy (**b**). Correlation coefficients are displayed within each cell, with genes and Banff lesions hierarchically clustered according to their correlation patterns. **c,** Concordance of gene–gene correlation structures between BTM and biopsy, assessed by the Mantel test. **d,** Agreement of gene–lesion correlations between BTM and biopsy, assessed by Pearson correlation.

We further assessed the concordance between BTM- and biopsy-derived gene-expression profiles. Gene-gene distance structures were highly concordant between the two compartments, as demonstrated by Mantel analysis (Mantel *r* = 0.69, *P* < 0.001) (Figure 2c). Similarly, gene–lesion correlation patterns showed strong agreement between BTM and biopsy tissues (*r* = 0.67, *P* < 0.001) (Figure 2d), indicating that the overall relationships between immune-related gene expression and Banff histologic lesions were largely preserved in the BTM.

### BTM gene-panel models

#### Shared and outcome-specific predictive features of BTM gene-panel models

The features contributing most strongly to the best-tuned machine-learning models are shown in Supplementary Figure 2. Across both binary rejection classification and continuous outcome prediction, a broadly consistent set of genes emerged as important features despite differences among the three machine-learning algorithms. *GBP1, CXCL10, CD8A, GNLY, CTLA4, SH2D1B, VCAM1,* and *MS4A7* were consistently ranked among the most influential features. *GBP1* and *CXCL10* showed the greatest importance for rejection classification, whereas *CXCL10, CD8A, CTLA4,* and *GBP1* were frequently identified as key features for predicting the AMR/MVI, TCMR/TI, and Activity indices. *TPSAB1, PECAM1,* and *VCAM1* showed greater importance for prediction of the Chronicity Index, suggesting a distinct feature profile for chronic versus active allograft injury.

#### Predictive performance for rejection

The performance distributions of the best-tuned models are shown in Figure 3, with corresponding summary metrics presented in Supplementary Table 3. All three machine-learning algorithms demonstrated stable predictive performance, with median AUCs ranging from 0.75 to 0.80. EN achieved the highest median AUC (0.80 [0.75, 0.87]), with SVM-RBF showing closely comparable performance. ROC curves largely overlapped across the three algorithms, indicating similar discriminatory performance. Modest differences were observed in probability calibration, with EN demonstrating the best overall calibration (slope, 0.92 [0.87, 0.98]; intercept, 0.08 [0.01, 0.10]; Brier score, 0.16 [0.15, 0.16]). Decision curve analysis demonstrated that all three models provided a positive net benefit compared with treat-all (calling every biopsy a rejection *a priori*, i.e. blindly treating all patients as if they would have a rejection) and treat-none (calling all biopsies free of rejection *a priori*, i.e. no anti-rejection therapy) strategies across a broad range of threshold probabilities. EN and SVM-RBF provided modestly greater net benefit than RF across intermediate threshold probabilities.

**Figure 3.**
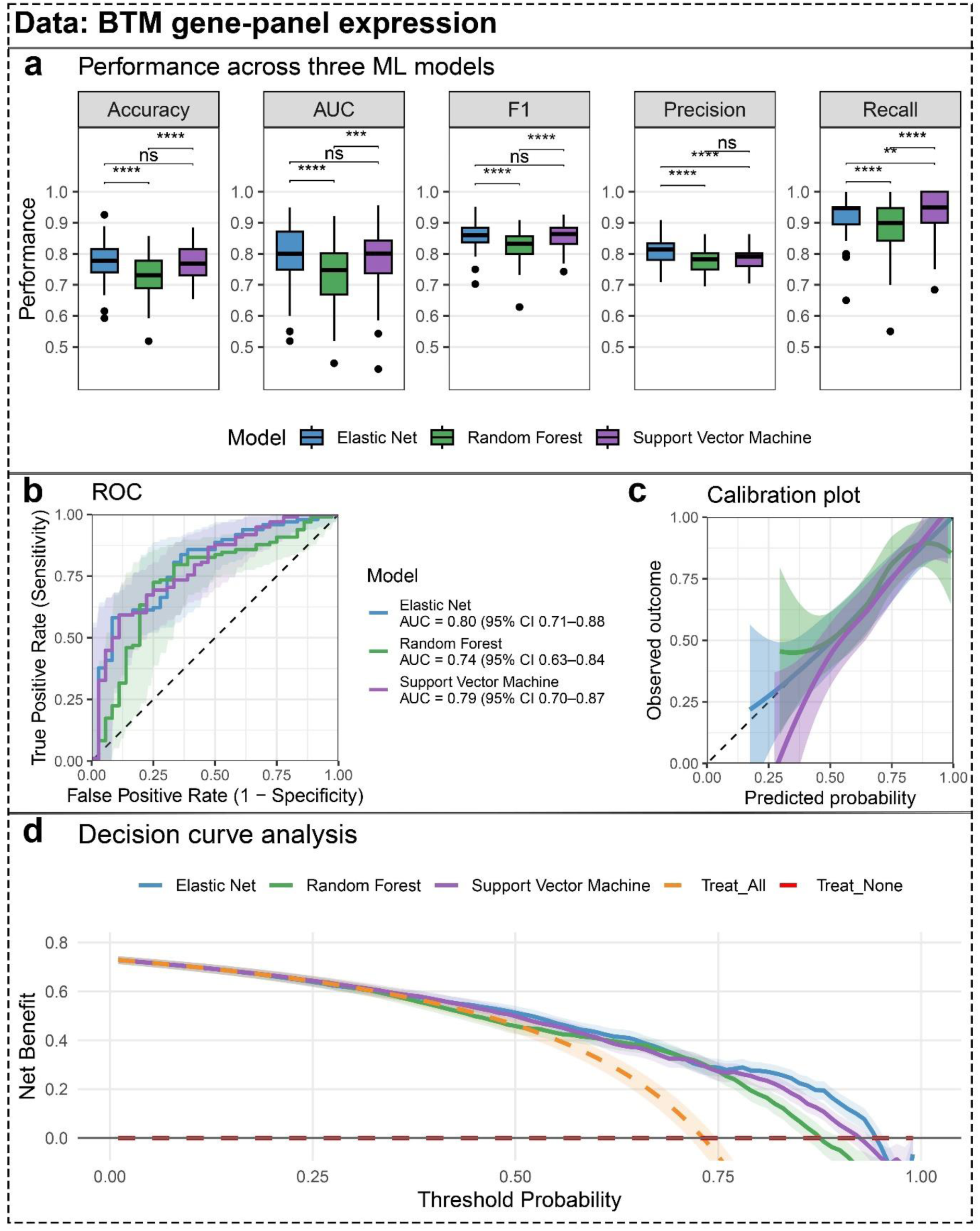
Performance of machine-learning models for binary diagnostic classification using BTM gene-panel expression. a, Comparison of classification performance across Elastic Net, Random Forest, and SVM-RBF models, evaluated by accuracy, AUC, F1 score, precision, and recall. Model performance was evaluated using 20 repetitions of five-fold cross-validation. Within each repetition, out-of-fold predictions from the five held-out folds were combined to obtain a single performance estimate, yielding 20 matched estimates per model. Pairwise differences between models were assessed using paired Wilcoxon signed-rank tests across matched repetitions. Statistical significance is indicated above the pairwise comparisons; ns, not significant. **b,** Out-of-fold predicted probabilities were averaged across cross-validation repetitions for each patient, and the resulting patient-level probabilities were used to construct the ROC curves. Shaded regions represent pointwise 95% confidence bands for the ROC curves, estimated using bootstrap resampling. AUCs are reported with 95% confidence intervals calculated using DeLong’s method. **c,** Calibration curves showing the relationship between the averaged patient-level out-of-fold predicted probabilities and observed outcomes for each model. The dashed diagonal line indicates perfect calibration, and shaded areas represent pointwise 95% confidence intervals around the smoothed calibration curves. **d,** Decision curve analysis showing the net benefit of the Elastic Net, Random Forest, and SVM models across a range of threshold probabilities. The dashed orange and red lines indicate the strategies of treating all patients (calling every biopsy a rejection a priori, i.e. blindly treating all patients as if they would have a rejection) and treating no patients (calling all biopsies free of rejection a priori, i.e. no anti-rejection therapy), respectively.

#### Predictive performance for Continuous Indices

Predictive performance varied across the four continuous indices (Figure 4 and Supplementary Table 4). For the Activity Index, EN outperformed the other algorithms, achieving the highest R² (0.33) and lowest RMSE (3.21). Performance was lower for the Chronicity Index across all three algorithms, although EN showed the best overall performance (R² = 0.12; RMSE = 3.20). For the AMR/MVI Index, differences among algorithms were modest, with RF achieving the highest R² (0.27) and lowest RMSE (2.74). For the TCMR/TI Index, SVM-RBF demonstrated the best overall performance, with the highest R² (0.31) and lowest RMSE (1.79).

**Figure 4.**
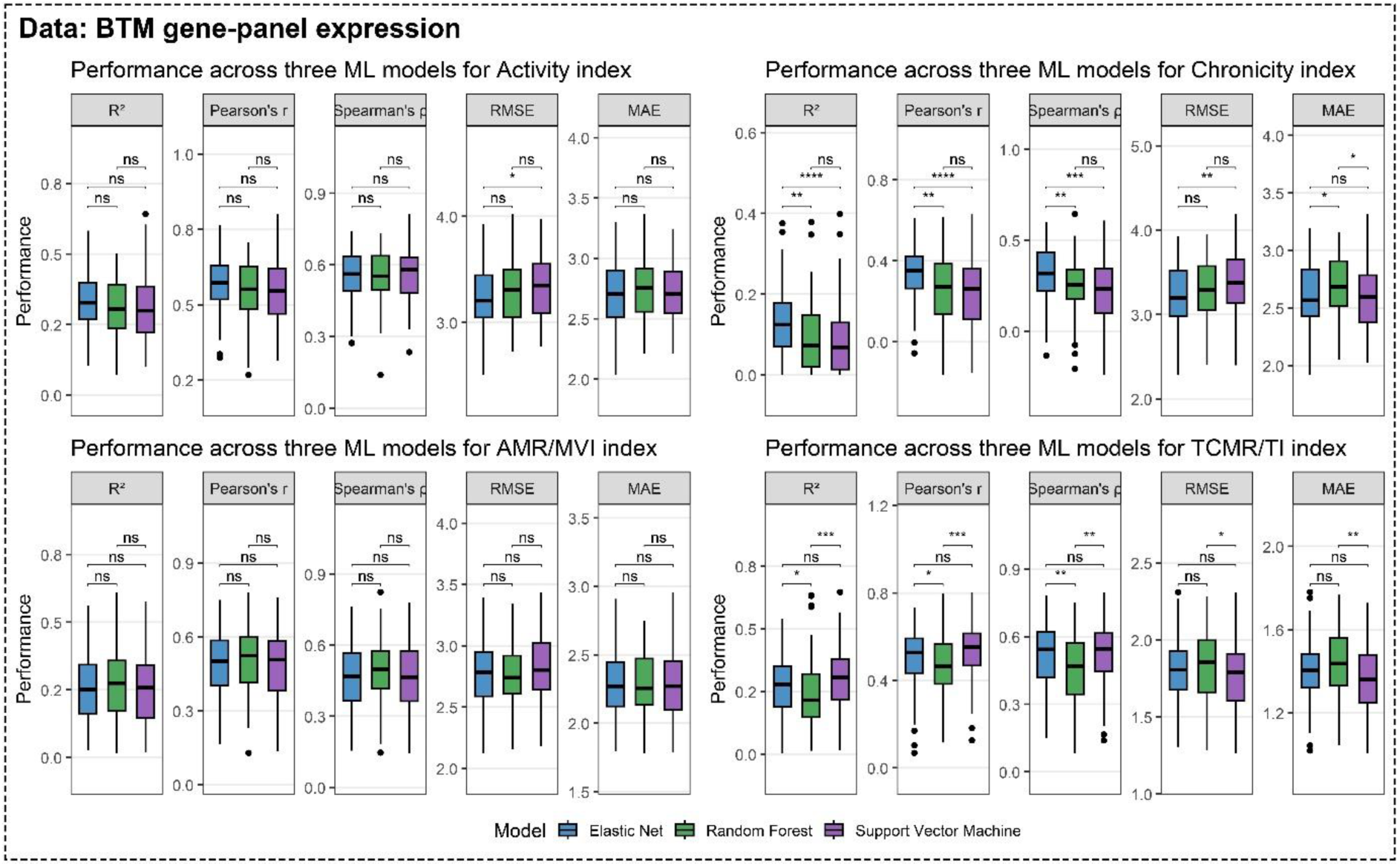
Performance of machine-learning models for continuous index prediction using BTM gene-panel expression. Prediction performance of Elastic Net, Random Forest, and SVM-RBF models for the Activity, Chronicity, AMR/MVI, and TCMR/TI indices. Model performance was evaluated using 20 repetitions of five-fold cross-validation. Within each repetition, out-of-fold predictions from the five held-out folds were combined to obtain a single performance estimate, yielding 20 matched estimates per model. Predictive performance was assessed using the coefficient of determination (R²), Pearson’s correlation coefficient (*r*), Spearman’s rank correlation coefficient (ρ), root mean squared error (RMSE), and mean absolute error (MAE). Boxplots show the distributions of performance estimates across the 20 cross-validation repetitions. Pairwise differences between models were assessed across matched repetitions using paired Wilcoxon signed-rank tests. Statistical significance is indicated above the pairwise comparisons; ns, not significant.

### Comparison of BTM gene-panel, biopsy gene-panel, and biopsy whole-transcript models

#### Predictive performance across modeling datasets

Predictive performance varied across the three modeling datasets, with the magnitude of between-dataset differences depending on the outcome. Pairwise differences (Δ) in performance metrics across the three modeling datasets using EN algorithm are summarized in Figure 5 and Supplementary Table 5.

**Figure 5.**
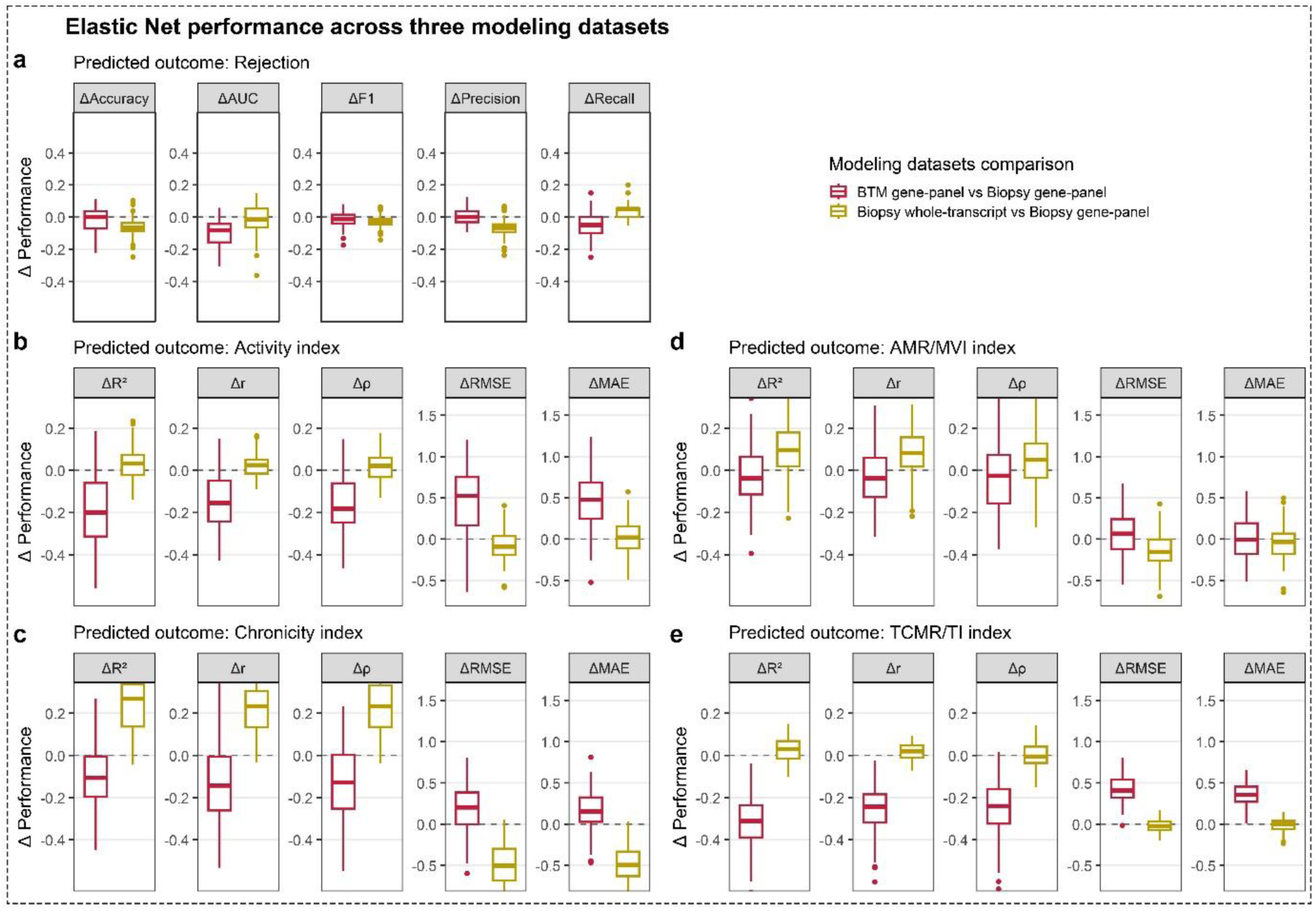
Comparison of Elastic Net predictive performance across modeling datasets. Differences in Elastic Net model performance using the BTM gene panel versus the biopsy gene panel (red) and the biopsy whole-transcriptome versus the biopsy gene panel (yellow). Performance differences (Δ) were calculated relative to models based on the biopsy gene panel, such that positive values indicate improved performance relative to the biopsy gene-panel model and negative values indicate reduced performance. **a,** Differences in binary diagnostic classification performance for rejection, evaluated by Δ accuracy, Δ AUC, Δ F1 score, Δ precision, and Δ recall. **b–e,** Differences in continuous prediction performance for the Activity (**b**), Chronicity (**c**), AMR/MVI (**d**), and TCMR/TI (**e**) indices, evaluated by the coefficient of determination (ΔR²), Pearson’s correlation coefficient (Δ*r*), Spearman’s rank correlation coefficient (Δρ), root mean squared error (ΔRMSE), and mean absolute error (ΔMAE). Models were evaluated using 20 repetitions of five-fold cross-validation, with performance differences calculated across matched cross-validation repetitions. Boxplots show the distributions of performance differences across the 20 repetitions.

For binary rejection classification, BTM gene-panel models did not show a great inferiority discrimination than biopsy gene-panel models, with a ΔAUC of −0.08 (−0.16, −0.04) and a ΔF1 of −0.01 (−0.04, 0.01). Compared with biopsy gene-panel models, biopsy whole-transcript models showed a ΔAUC of −0.02 (−0.06, 0.05) and a ΔF1 of −0.03 (−0.04, −0.01). Overall, BTM gene-panel models showed slightly lower performance than biopsy gene-panel models, whereas expanding from the predefined gene panel to the whole transcriptome in biopsy samples did not improve binary classification performance.

For prediction of Continuous Indices, differences between BTM gene-panel and biopsy gene-panel models varied across the four indices. The smallest differences were observed for the AMR/MVI Index (ΔR² = −0.04; ΔRMSE = 0.07), indicating broadly comparable performance for this targeted panel. Larger differences were observed for the Activity and Chronicity Indices, with ΔR² values of −0.20 and −0.11 and ΔRMSE values of 0.52 and 0.20, respectively. The largest difference was observed for the TCMR/TI Index (ΔR² = −0.31; ΔRMSE = 0.41). In contrast, biopsy whole-transcript and biopsy gene-panel models showed broadly comparable performance for continuous index prediction. For the Activity, AMR/MVI, and TCMR/TI Indices, median ΔR² ranged from 0.01 to 0.30 and median ΔRMSE from −0.16 to −0.02, indicating only minor differences between the two modeling datasets. The Chronicity Index was an exception, for which the biopsy whole-transcript model showed better performance (ΔR² = 0.27; ΔRMSE = −0.50).

#### Predefined gene panel versus transcriptome-wide feature selection

We next assessed whether transcriptome-wide feature selection improved predictive performance beyond that achieved with the predefined gene panel (Figure 6). For binary rejection classification, the optimal transcriptome-wide models retained 69, 250, and 19 genes for EN, RF, and SVM-RBF, respectively, without substantial improvement over the predefined 19-gene panel. A similar pattern was observed for continuous outcomes except Chronicity Index, with larger feature sets generally providing little or no improvement in predictive performance. RF showed modest improvements for the Activity and AMR/MVI Indices using 60 genes, while retaining 19 genes for the Chronicity Index. For the TCMR/TI Index, a 19-gene RF model achieved a modest improvement in R² (ΔR² = 0.10) relative to the predefined panel. Overall, transcriptome-wide feature selection increased model complexity without yielding consistent improvements in predictive performance, indicating that further selection of genes for BTM profiling likely would not add extra discriminatory value.

**Figure 6.**
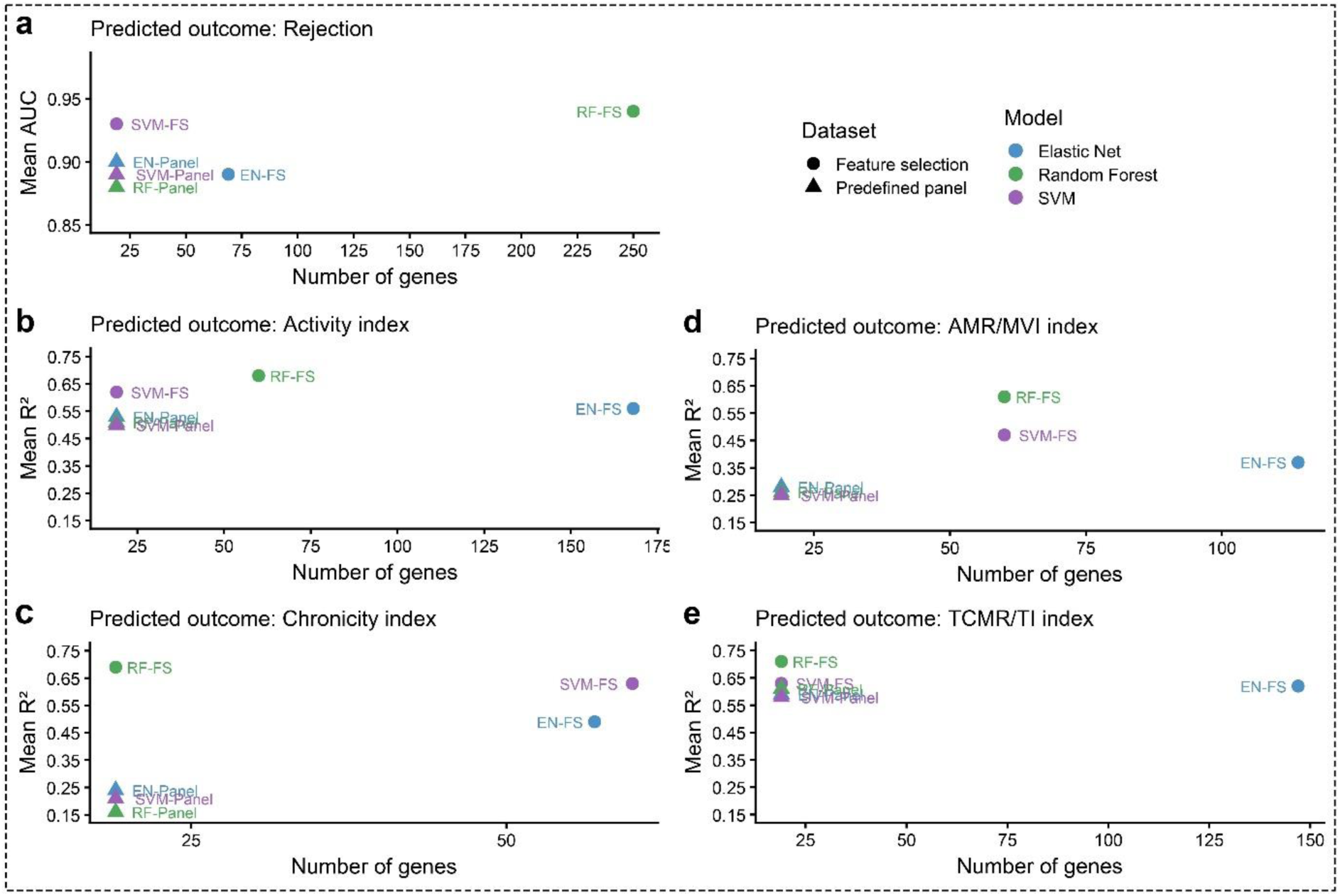
Trade-off between predictive performance and gene-panel size across machine-learning models. Predictive performance is plotted against the number of genes included in each model for Elastic Net (EN), Random Forest (RF), and Support Vector Machine (SVM). Models based on predefined gene panels are indicated by triangles, whereas models using feature-selected gene sets (FS) are indicated by circles. Colors denote the machine-learning algorithms. **a,** Performance for binary classification of rejection, assessed by the mean AUC. **b–e,** Performance for continuous prediction of the Activity (**b**), Chronicity (**c**), AMR/MVI (**d**), and TCMR/TI (**e**) indices, assessed by the mean coefficient of determination (R²). Mean performance was calculated across 20 repetitions of five-fold cross-validation. The x-axis indicates the number of genes included in each model.

## Discussion

In this study, we propose an innovative molecular diagnostic workflow that combines qPCR-based measurement of a 19-gene panel in BTM with gene-expression models for the molecular assessment of kidney transplant biopsies. This approach leverages qPCR, a widely available, rapid, robust, and cost-effective technology, together with BTM as a readily accessible source of biopsy-derived transcripts, with the potential to facilitate rapid molecular assessment without relying on additional biopsy tissue. We evaluated an 19-gene panel using two transcriptomic sources derived from the same biopsy: BTM and the corresponding FFPE tissue, which served as the reference. As expected, the biopsy gene-panel models demonstrated excellent performance in both binary diagnostic classification and continuous indices prediction, supporting the feasibility of a small gene panel as a practical approach for molecular diagnosis. Although the predictive performance of BTM gene-panel models was slightly lower than that of biopsy gene-panel models, they nevertheless achieved good discriminative ability. These findings demonstrate that BTM can be purposed as a rapid and specific transcriptomic source for molecular diagnosis using a simple qPCR assay.

Molecular diagnosis has become an increasingly important component of post-transplant management. Instead of replacing conventional histopathology, transcriptomic analysis provides complementary biological information that improves the understanding of heterogeneous rejection phenotypes and may facilitate more personalized therapeutic strategies. Current molecular diagnostic approaches, including the Molecular Microscope Diagnostic System (MMDx), the B-HOT NanoString platform, and bulk RNA sequencing, have demonstrated the clinical value of gene-expression profiling in kidney transplantation. However, their broader implementation remains limited by the need for specialized platforms, relatively high costs, prolonged turnaround times, and dependence on biopsy tissue for molecular analysis^10,19–21^. The proposed BTM–qPCR workflow addresses several of these limitations. As a by-product generated during routine biopsy processing, BTM represents an untapped transcriptomic resource that requires no additional tissue sampling. Combined with qPCR, a rapid, inexpensive, and widely available technology, this approach enables same-day molecular assessment using standard laboratory infrastructure. Rather than competing with comprehensive transcriptomic platforms, the BTM–qPCR workflow provides a pragmatic strategy for rapid molecular screening that may complement conventional pathology while more comprehensive histopathological evaluation is still in progress.

The greatest potential clinical utility of the BTM-qPCR workflow lies in the interval between biopsy acquisition and final histopathological interpretation, when treatment decisions may need to be made before a complete pathology report is available. For example, when a biopsy is submitted to the pathology laboratory on a Friday, a complete histopathological assessment may not be available until after the weekend, and the turnaround time may be further prolonged when additional staining, ancillary testing, or expert review is required. In this setting, BTM provides a readily available source of biopsy-derived transcripts that could generate molecular information shortly after biopsy acquisition without altering the routine biopsy workflow or consuming additional biopsy tissue. Consistent with this proposed application, decision curve analysis^22^ demonstrated that all three BTM-based prediction models provided a positive net benefit across a broad range of threshold probabilities compared with both treat-all and treat-none strategies. Thus, the BTM-qPCR workflow could complement histopathological assessment by providing an early indication of rejection probability while the final pathology diagnosis is pending, potentially facilitating prompt targeted intervention in patients at high risk of rejection while avoiding unnecessary intensification of immunosuppression in those at low risk.

Rapid identification of rejection has important clinical implications throughout the post-transplant course. Binary rejection classification represents the first step in clinical assessment, while continuous indices provide a more precise and granular assessment of inflammatory activity, lesion severity, and chronic tissue injury^15^. Accordingly, we evaluated whether the proposed BTM-qPCR workflow could predict continuous histopathological indices. Consistent with the biopsy gene-panel models, the BTM gene-panel models showed promising performance for the AMR/MVI, TCMR/TI, and Activity Indices, with the highest predictive accuracy observed for the Activity index. Prediction of the Chronicity Index was less accurate for both BTM gene panel and biopsy gene panel models. This finding is likely attributable to the design of our gene panel, which prioritized immune-related genes associated with rejection and robustly detectable in BTM based on our previous study^13^. In addition, BTM preferentially capture cells that more easily dissociate from the tissue, such as infiltrating immune cells and injured or detached endothelial and epithelial cells, whereas structurally embedded cells involved in fibrosis, including collagen-producing fibroblasts, may be less readily released from the biopsy tissue. Notably, *TPSAB1*, a mast cell marker, contributed to Chronicity Index prediction and showed strong correlations with Banff IFTA, i-IFTA, and t-IFTA lesions, suggesting that mast cell–associated signals in BTM may nevertheless provide molecular information related to chronic tissue injury. This is in line with the function of mast cells in the process of fibrogenesis as has been proposed in the literature^23–25^.

This study has several limitations. First, it was conducted at a single center, therefore, external validation in larger multicenter cohorts is an important next step. Second, although the predefined qPCR panel showed promising performance, transcriptome-wide feature selection identified additional genes with strong predictive signals for specific Continuous Indices. Further investigation of these candidate genes in BTM may help optimize the current gene panel and improve its predictive performance.

In conclusion, our findings support the feasibility of using BTM-derived gene expression for molecular assessment of kidney transplant biopsies. Despite slightly lower predictive performance than biopsy tissue, BTM provided informative molecular signals for both rejection classification and continuous indices. These results establish a basis for further validation and refinement of the BTM–qPCR approach as a complementary tool to support timely and integrated assessment of kidney allograft pathology.

## Data Availability

All data produced in the present study are available upon reasonable request to the authors

## Data Sharing Statement

The data that support the findings of this study are available from the principal investigator of this study (JK) at upon reasonable request, subject to institutional, ethical and legal restrictions.

## Acknowledgments

The study is supported by the China Scholarship Council.

---

**Supplementary Table 1.**
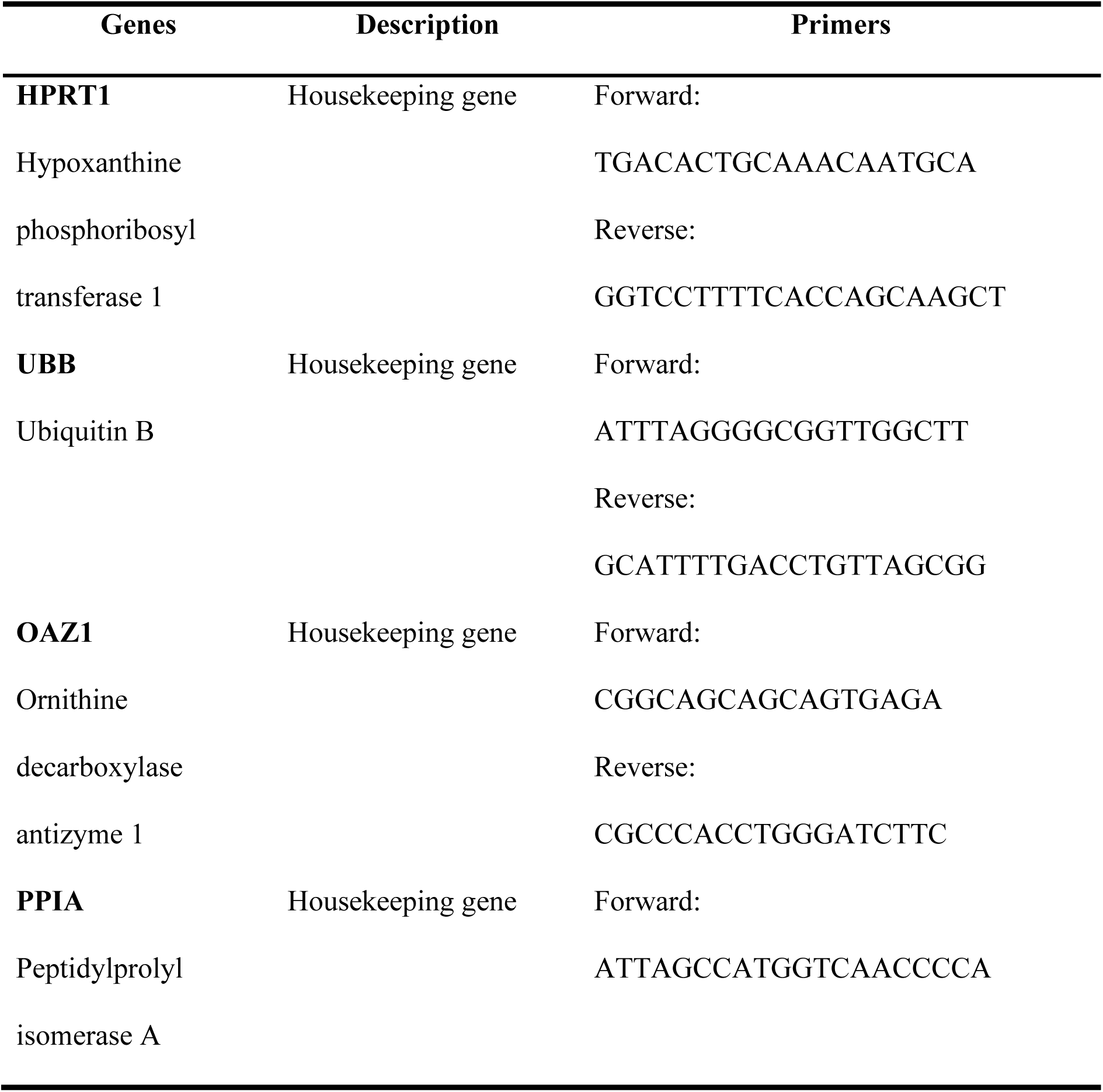
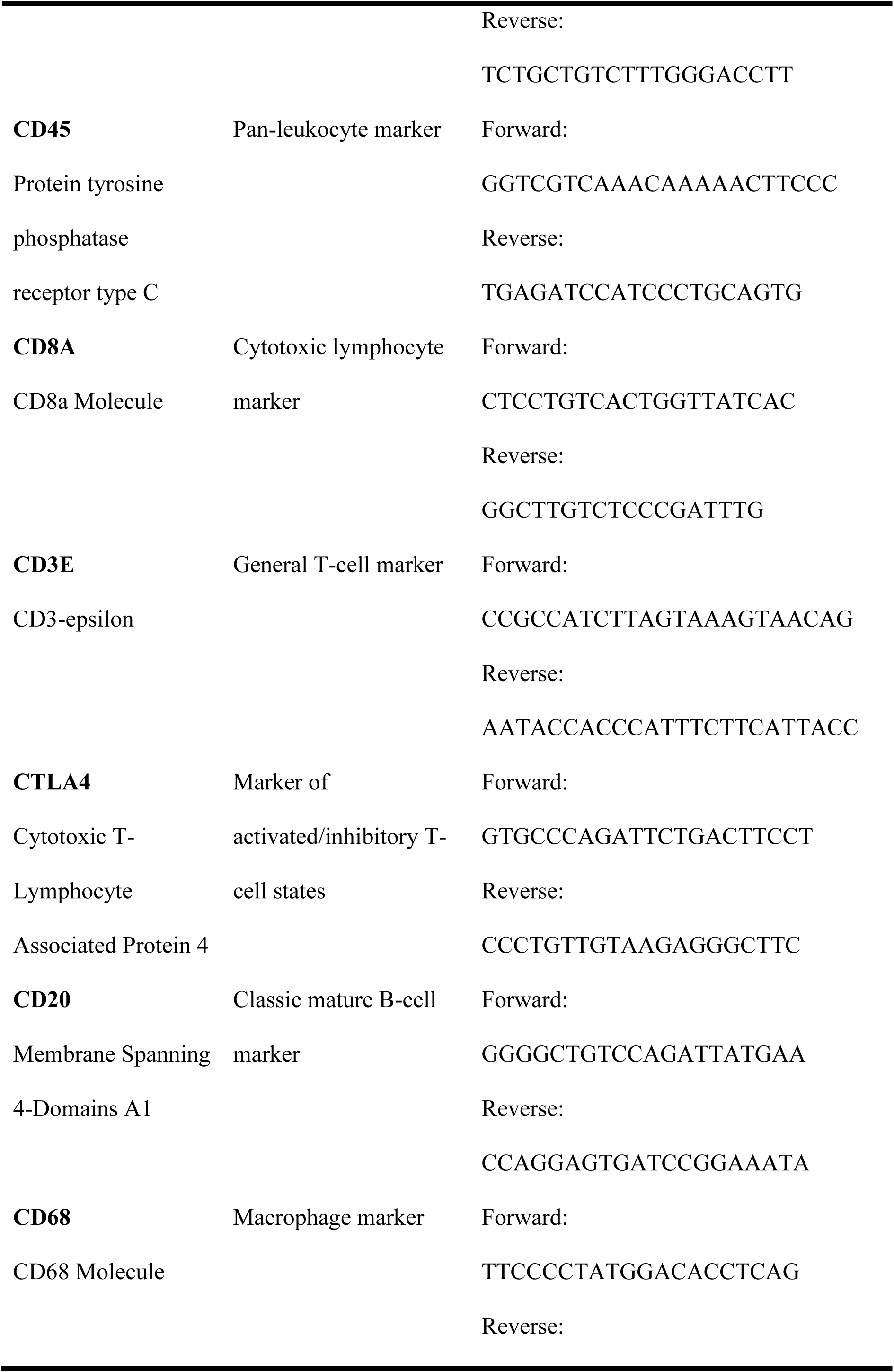
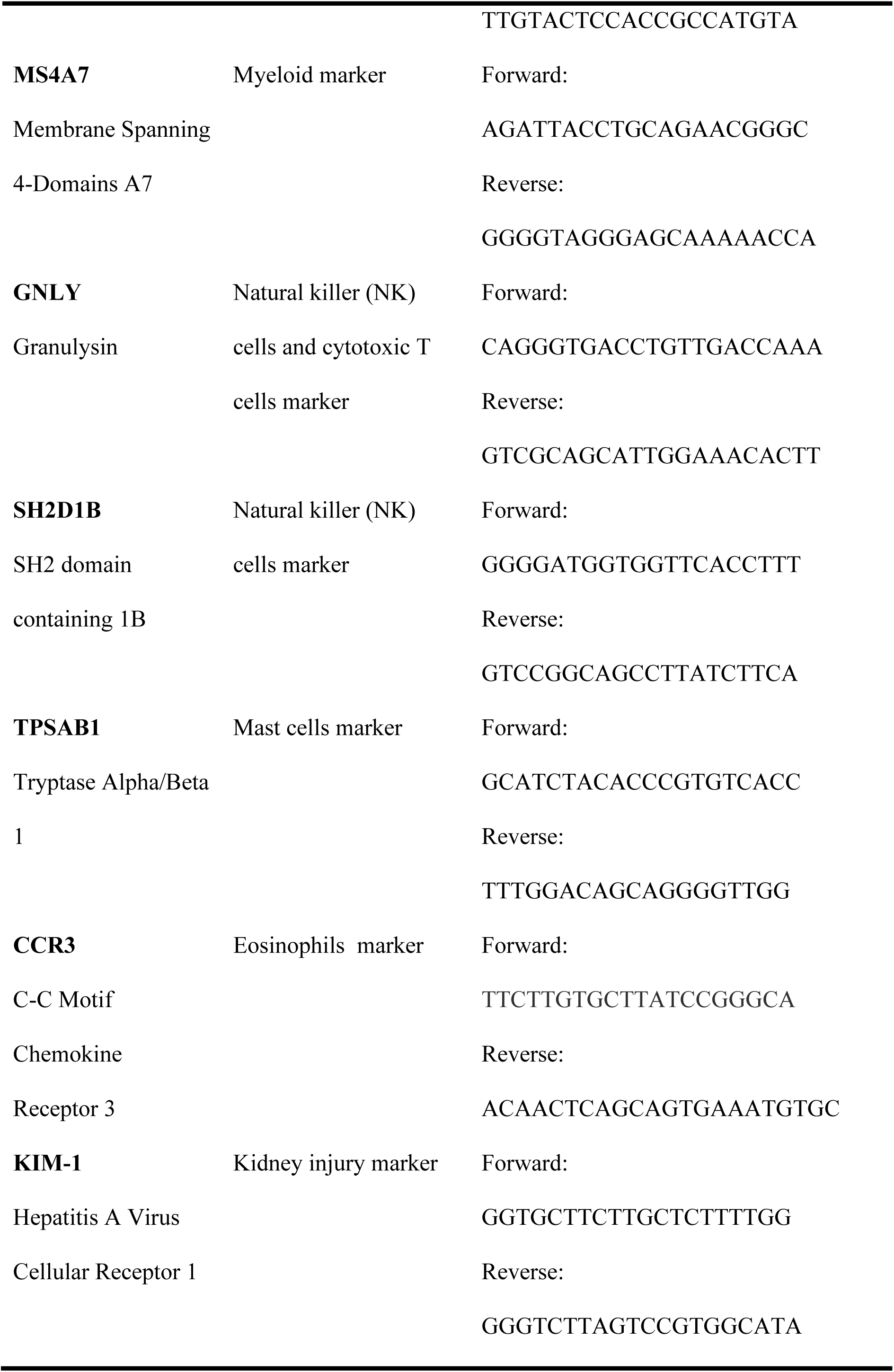
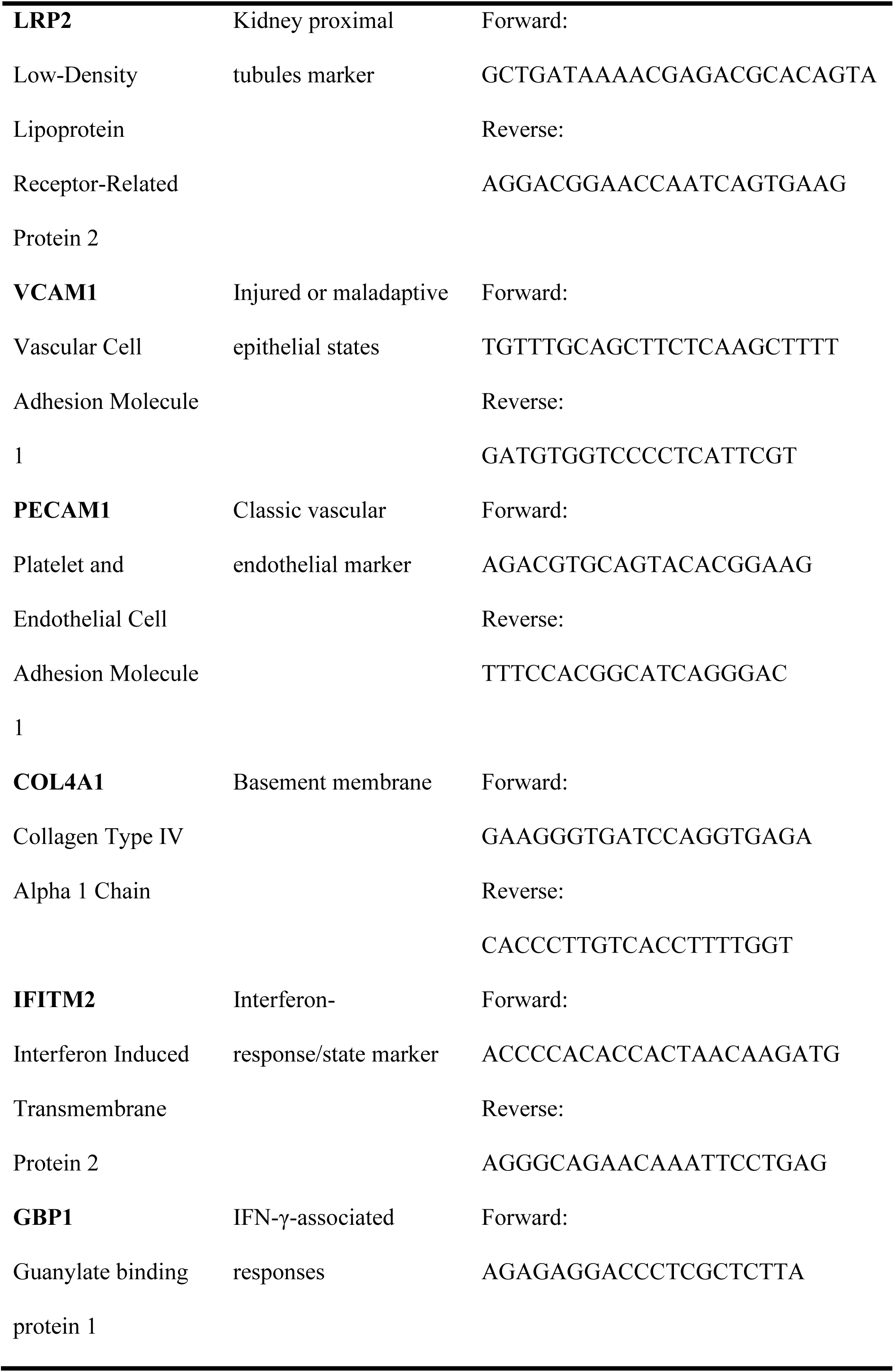
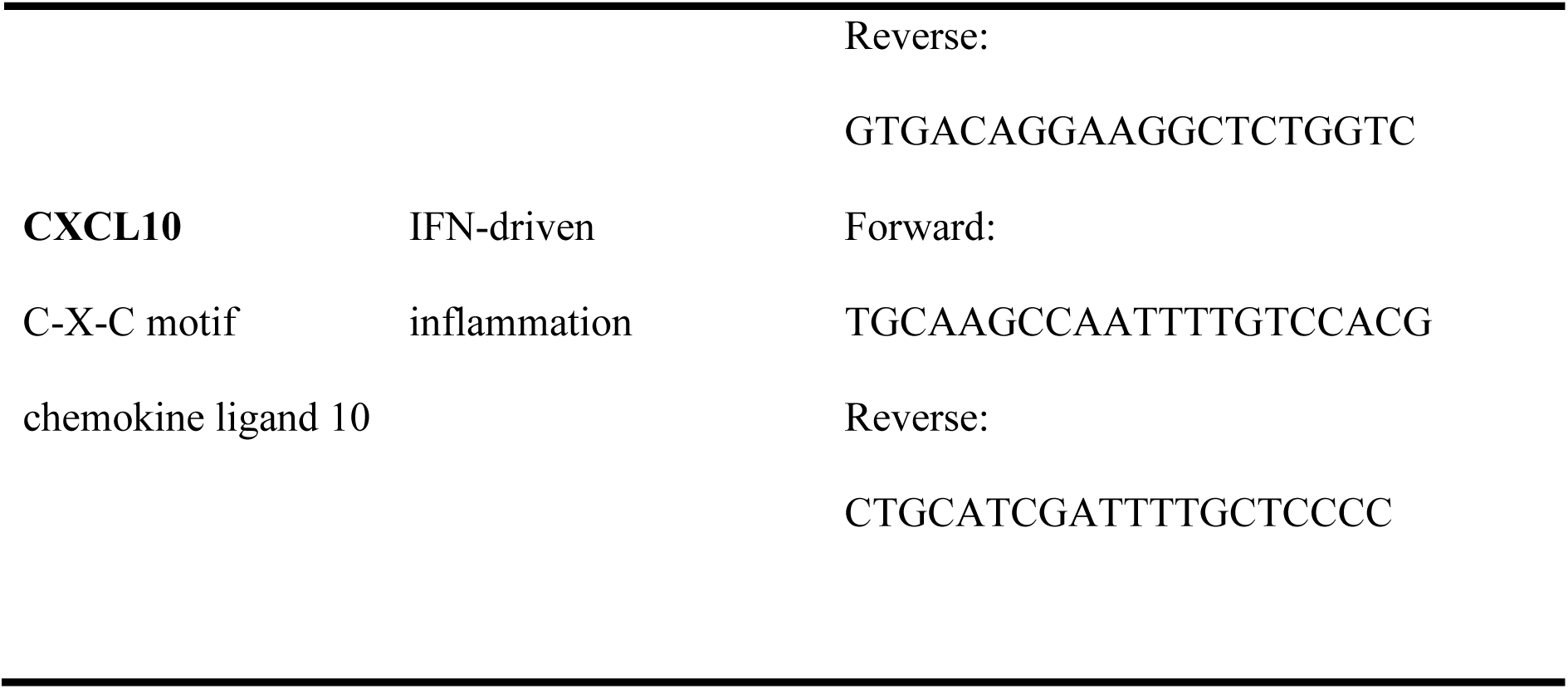
Genes and primer sequences analyzed in PCR experiments.

**Supplementary Table 2.** Histological characteristics in the study population.

| Variable | N | N = 157 |
| --- | --- | --- |
| glom | 134 | 22 (14, 30) |
| fsgs | 134 |  |
| absent |  | 101 (75%) |
| present |  | 33 (25%) |
| mpgn | 134 |  |
| absent |  | 129 (96%) |
| present |  | 5 (3.7%) |
| cresc | 134 |  |
| absent |  | 130 (97%) |
| present |  | 4 (3.0%) |
| tma | 134 | 25 (19%) |
| atn | 134 | 106 (79%) |
| g | 134 | 0 (0, 2) |
| cg | 134 | 0 (0, 0) |
| mm | 134 | 0 (0, 0) |
| i | 134 | 1 (0, 2) |
| t | 134 | 1 (0, 2) |
| ti | 134 | 2 (1, 3) |
| ifta | 134 | 1 (0, 2) |
| ci | 134 | 1 (0, 2) |
| ct | 134 | 1 (1, 2) |
| iifta | 134 | 1 (0, 3) |
| tifta | 134 | 0 (0, 1) |
| ptc | 134 | 1 (0, 2) |
| v | 129 | 0 (0, 0) |
| no arteries present |  | 5 |
| cv | 129 | 1 (0, 1) |
| no arteries present |  | 5 |
| ah | 134 | 1 (0, 1) |
| c4d positive | 134 | 57 (43%) |
| DSA positive | 134 |  |
| Negative |  | 98 (73%) |
| Positive |  | 36 (27%) |
| bk | 134 |  |
| negative |  | 49 (37%) |
| not performed |  | 83 (62%) |
| positive |  | 2 (1.5%) |

**Supplementary Table 3.** Comparison of predictive performance of the optimally tuned models for predicting rejection: BTM gene-panel expression.

| **Metric** | **Elastic Net** | **Random Forest** | **Support Vector Machine** |
| --- | --- | --- | --- |
| **AUC** | 0.80 ± 0.09<br>0.80 [0.75–0.87] | 0.74 ± 0.10<br>0.75 [0.67–0.80] | 0.78 ± 0.09<br>0.80 [0.74–0.84] |
| **Accuracy** | 0.78 ± 0.06<br>0.78 [0.74–0.81] | 0.73 ± 0.06<br>0.73 [0.69–0.78] | 0.77 ± 0.05<br>0.77 [0.73–0.81] |
| **Precision** | 0.81 ± 0.05<br>0.81 [0.78–0.83] | 0.78 ± 0.04<br>0.78 [0.75–0.80] | 0.78 ± 0.04<br>0.79 [0.76–0.80] |
| **Recall** | 0.92 ± 0.07<br>0.95 [0.89–0.95] | 0.88 ± 0.08<br>0.90 [0.84–0.95] | 0.95 ± 0.06<br>0.95 [0.90–1.00] |
| **F1** | 0.86 ± 0.04<br>0.86 [0.84–0.88] | 0.82 ± 0.04<br>0.83 [0.80–0.86] | 0.86 ± 0.04<br>0.86 [0.83–0.88] |
| **Calibration intercept** | 0.08 ± 0.07<br>0.08 [0.01–0.10] | 0.32 ± 0.09<br>0.32 [0.25–0.39] | -0.16 ± 0.23<br>-0.17 [-0.40–0.04] |
| **Calibration slope** | 0.93 ± 0.09<br>0.92 [0.87–0.98] | 0.69 ± 0.11<br>0.72 [0.63–0.74] | 1.15 ± 0.24<br>1.13 [0.94–1.39] |
| **Brier score** | 0.15 ± 0.01<br>0.16 [0.15–0.16] | 0.17 ± 0.01<br>0.17 [0.17–0.17] | 0.16 ± 0.01<br>0.16 [0.15–0.16] |

**Supplementary Table 4.** Comparison of predictive performance of the optimally tuned models for predicting continuous index: BTM gene-panel expression.

| **Index** | **Metric** | **Elastic Net** | **Random Forest** | **Support Vector Machine** |
| --- | --- | --- | --- | --- |
| Activity | RMSE | 3.24 ± 0.30 | 3.30 ± 0.31 | 3.35 ± 0.31 |
|  |  | 3.21 [3.05–3.45] | 3.31 [3.05–3.49] | 3.35 [3.09–3.55] |
|  | MAE | 2.70 ± 0.27 | 2.74 ± 0.26 | 2.72 ± 0.24 |
|  |  | 2.71 [2.51–2.90] | 2.75 [2.56–2.91] | 2.71 [2.55–2.89] |

| Index | Metric | Elastic Net | Random Forest | Support Vector Machine |
| --- | --- | --- | --- | --- |
| | $R^2$ | $0.33 \pm 0.10$<br>0.33 [0.27–0.40] | $0.30 \pm 0.10$<br>0.30 [0.24–0.39] | $0.31 \pm 0.11$<br>0.30 [0.22–0.39] |
| | Pearson's r | $0.57 \pm 0.09$<br>0.57 [0.52–0.63] | $0.54 \pm 0.10$<br>0.55 [0.49–0.63] | $0.55 \pm 0.10$<br>0.55 [0.47–0.62] |
| | Spearman's $\rho$ | $0.57 \pm 0.11$<br>0.56 [0.49–0.64] | $0.55 \pm 0.11$<br>0.55 [0.50–0.64] | $0.56 \pm 0.11$<br>0.58 [0.48–0.63] |
| Chronicity | RMSE | $3.21 \pm 0.35$<br>3.20 [2.98–3.52] | $3.30 \pm 0.33$<br>3.29 [3.05–3.57] | $3.39 \pm 0.39$<br>3.38 [3.14–3.65] |
| | MAE | $2.61 \pm 0.26$<br>2.57 [2.43–2.84] | $2.69 \pm 0.26$<br>2.68 [2.52–2.91] | $2.60 \pm 0.27$<br>2.60 [2.37–2.78] |
| | $R^2$ | $0.13 \pm 0.09$<br>0.12 [0.07–0.18] | $0.09 \pm 0.08$<br>0.07 [0.02–0.15] | $0.08 \pm 0.08$<br>0.07 [0.01–0.13] |
| | Pearson's r | $0.34 \pm 0.14$<br>0.35 [0.26–0.42] | $0.27 \pm 0.15$<br>0.27 [0.14–0.38] | $0.24 \pm 0.17$<br>0.26 [0.11–0.36] |
| | Spearman's $\rho$ | $0.31 \pm 0.15$<br>0.32 [0.22–0.43] | $0.25 \pm 0.15$<br>0.26 [0.18–0.34] | $0.22 \pm 0.18$<br>0.23 [0.10–0.34] |
| AMR/MVI | RMSE | $2.77 \pm 0.25$<br>2.78 [2.59–2.95] | $2.76 \pm 0.24$<br>2.74 [2.61–2.92] | $2.82 \pm 0.25$<br>2.80 [2.64–3.02] |
| | MAE | $2.30 \pm 0.23$<br>2.27 [2.12–2.44] | $2.28 \pm 0.23$<br>2.26 [2.13–2.47] | $2.29 \pm 0.23$<br>2.27 [2.10–2.45] |
| | $R^2$ | $0.26 \pm 0.13$<br>0.25 [0.16–0.34] | $0.27 \pm 0.13$<br>0.27 [0.17–0.36] | $0.25 \pm 0.13$<br>0.26 [0.15–0.34] |
| | Pearson's r | $0.49 \pm 0.14$<br>0.50 [0.40–0.59] | $0.50 \pm 0.14$<br>0.52 [0.41–0.60] | $0.48 \pm 0.14$<br>0.51 [0.38–0.58] |
| | Spearman's $\rho$ | $0.47 \pm 0.14$<br>0.47 [0.36–0.57] | $0.49 \pm 0.13$<br>0.50 [0.42–0.57] | $0.46 \pm 0.14$<br>0.46 [0.36–0.58] |
| TCMR/TI | RMSE | $1.80 \pm 0.20$<br>1.81 [1.68–1.93] | $1.83 \pm 0.22$<br>1.85 [1.66–2.00] | $1.77 \pm 0.23$<br>1.79 [1.60–1.91] |
| | MAE | $1.41 \pm 0.15$<br>1.40 [1.32–1.48] | $1.44 \pm 0.16$<br>1.44 [1.33–1.56] | $1.37 \pm 0.16$<br>1.36 [1.25–1.48] |
| | $R^2$ | $0.27 \pm 0.12$ | $0.24 \pm 0.14$ | $0.30 \pm 0.13$ |
|  |  | 0.28 [0.19–0.35] | 0.22 [0.15–0.32] | 0.31 [0.22–0.38] |
| | Pearson's $r$ | $0.50 \pm 0.13$ | $0.47 \pm 0.15$ | $0.53 \pm 0.13$ |
|  |  | 0.53 [0.43–0.59] | 0.46 [0.38–0.57] | 0.55 [0.47–0.62] |
| | Spearman's $\rho$ | $0.52 \pm 0.14$ | $0.46 \pm 0.16$ | $0.52 \pm 0.14$ |
|  |  | 0.54 [0.42–0.62] | 0.47 [0.34–0.57] | 0.55 [0.44–0.62] |

**Supplementary Table 5.**
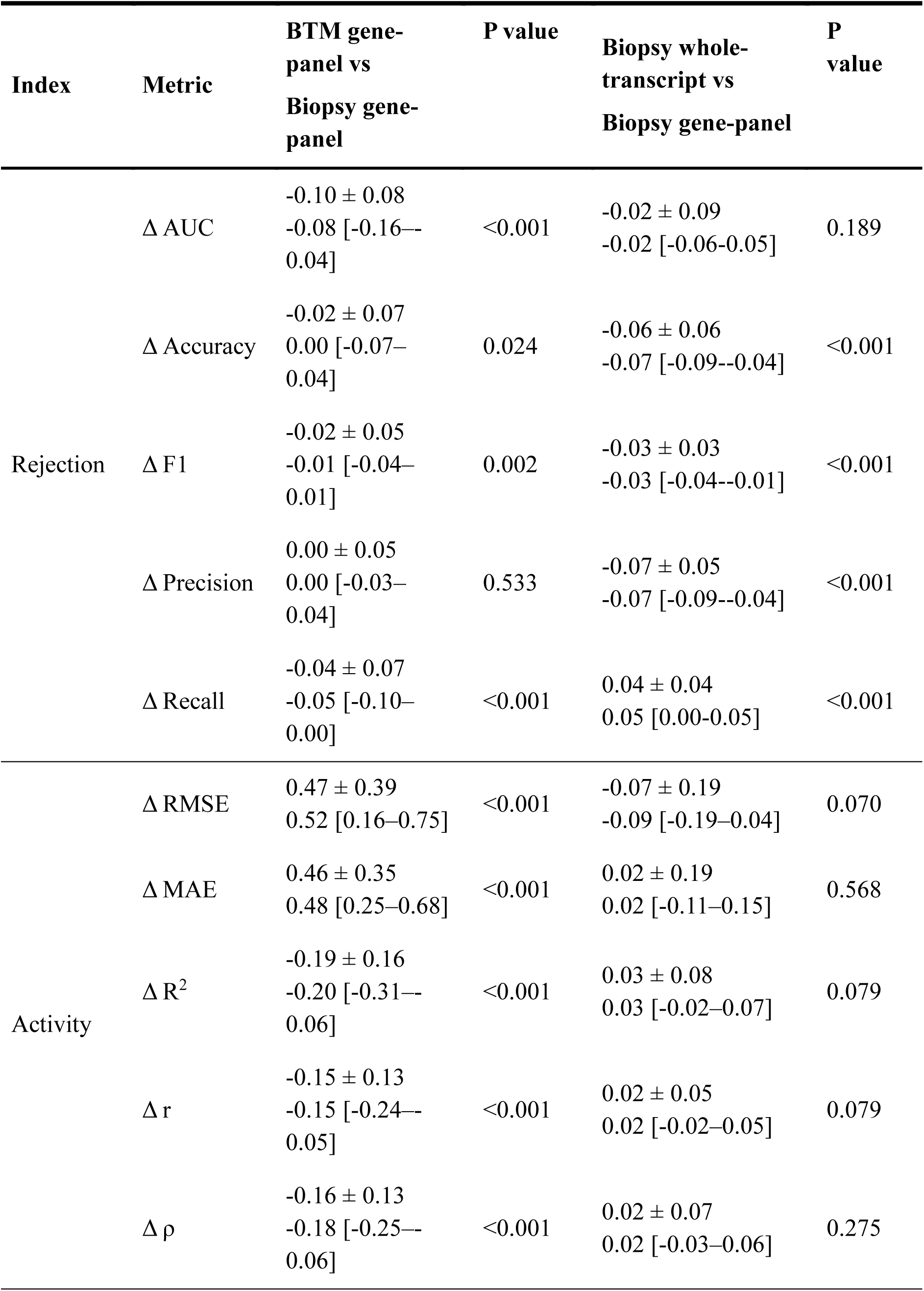
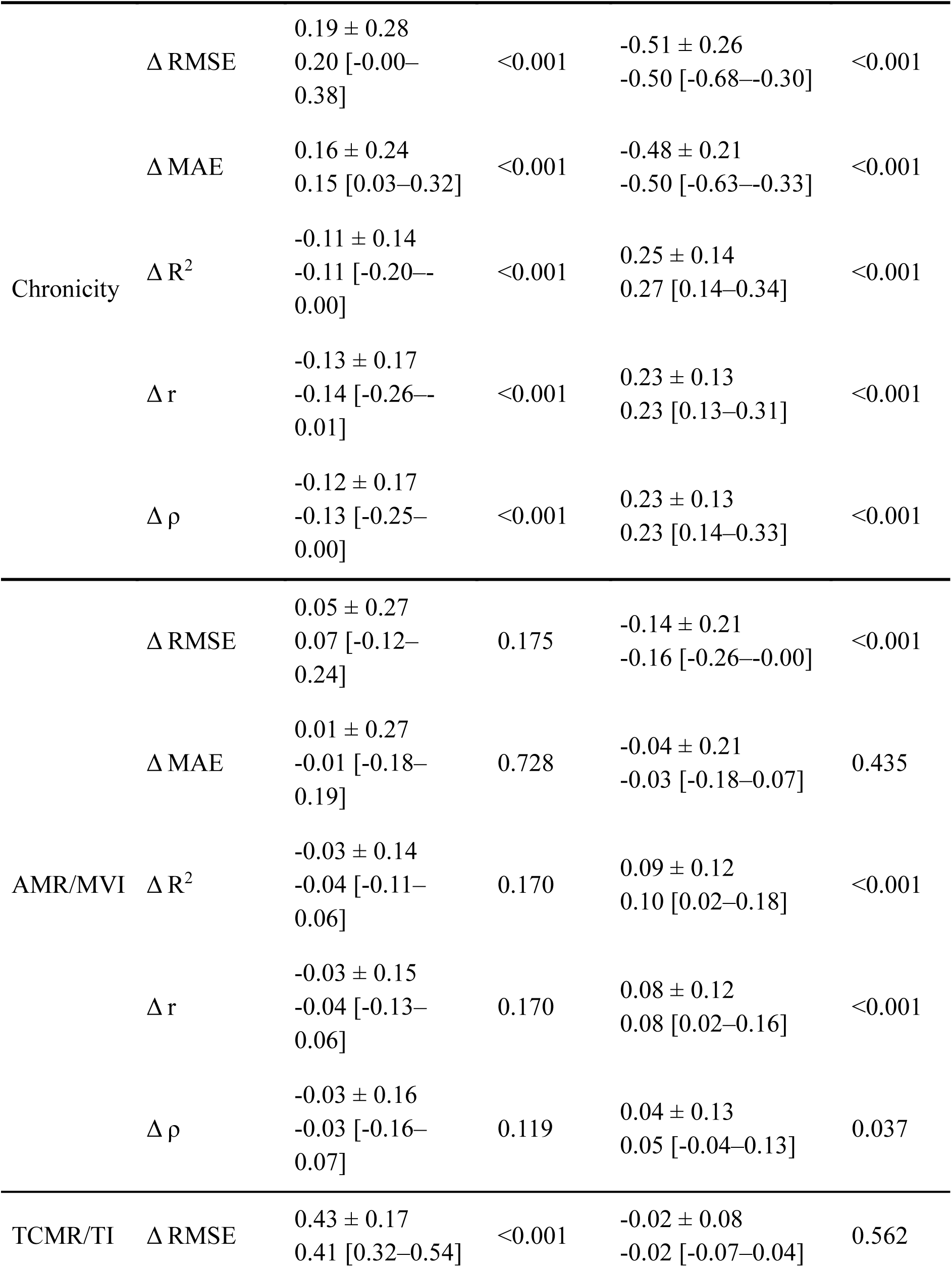
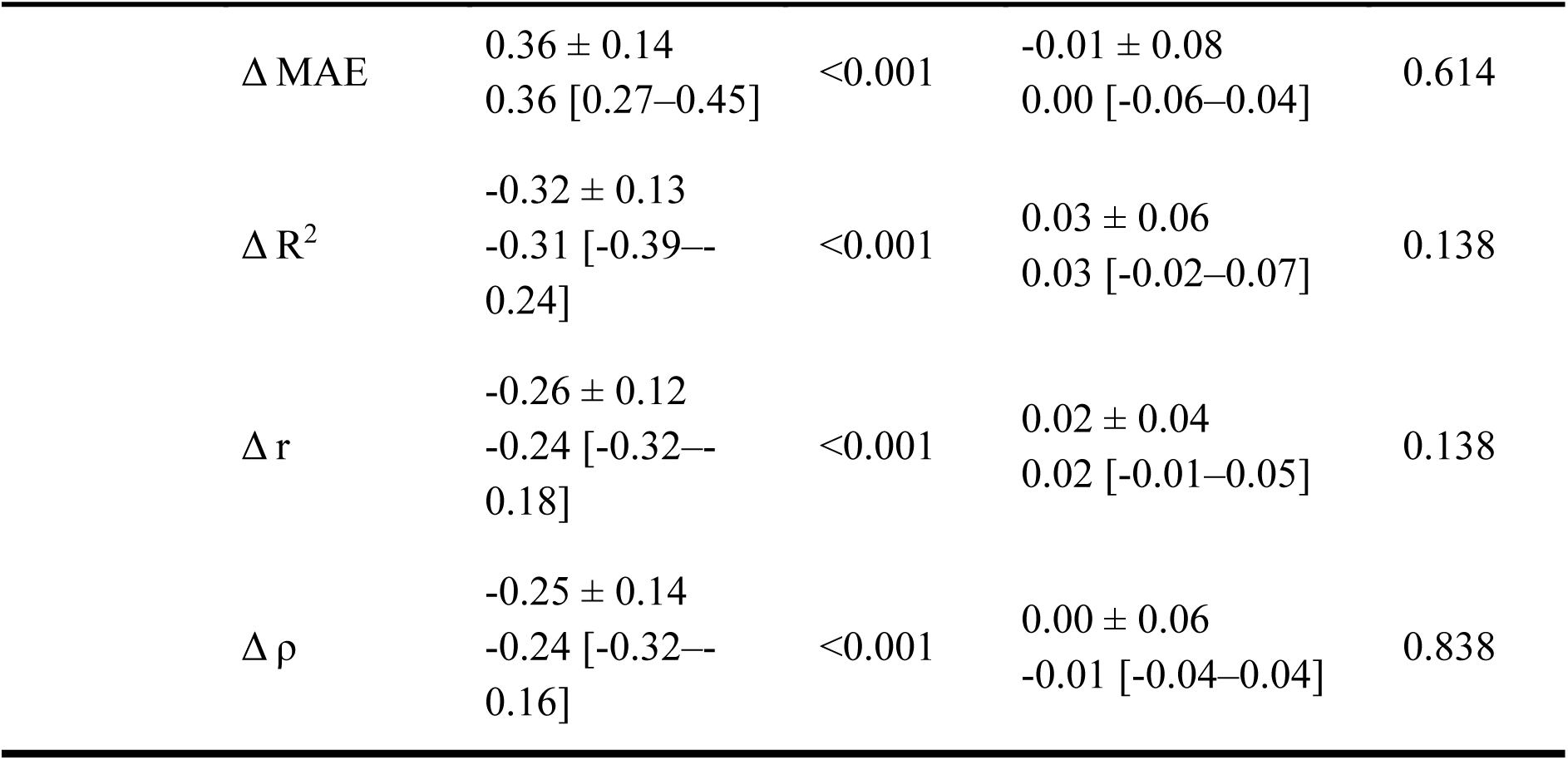
Pairwise differences (Δ) in predictive performance metrics across modeling datasets using the Elastic Net algorithm.

**Supplementary Table 6.**
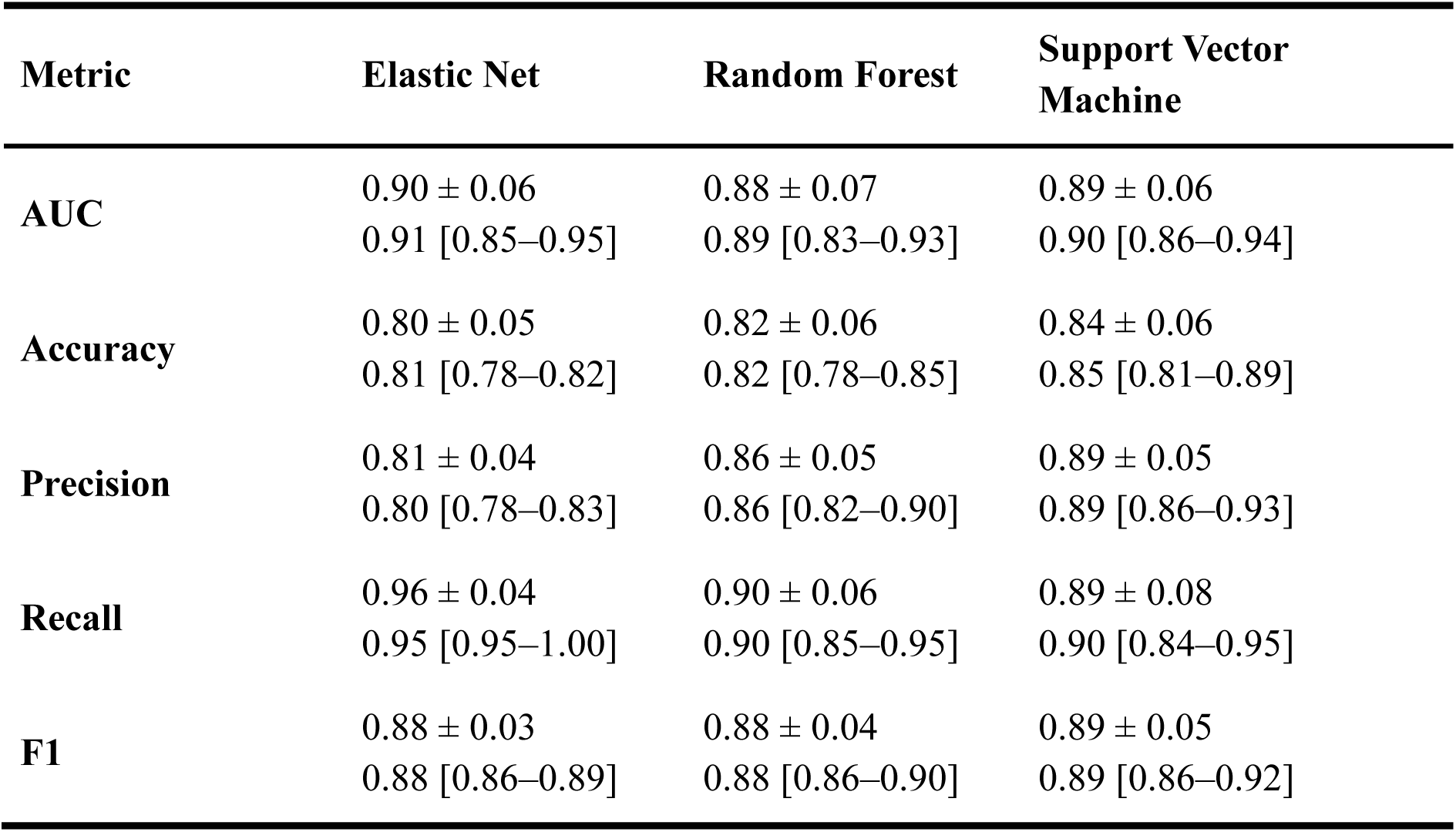
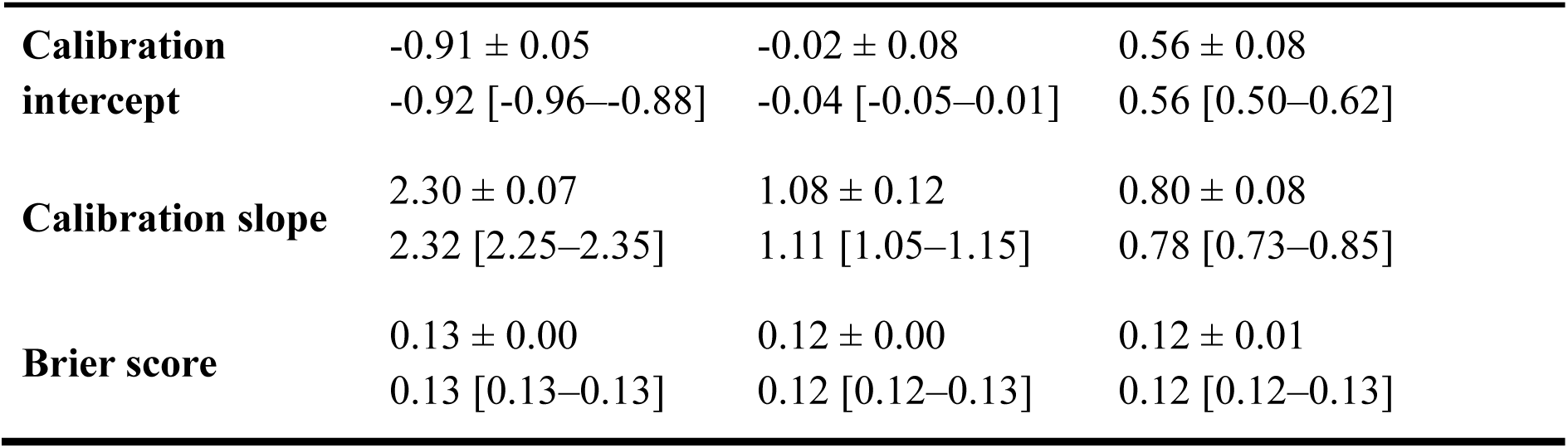
Comparison of predictive performance of the optimally tuned models for predicting rejection: Biopsy gene-panel expression.

**Supplementary Table 7.**
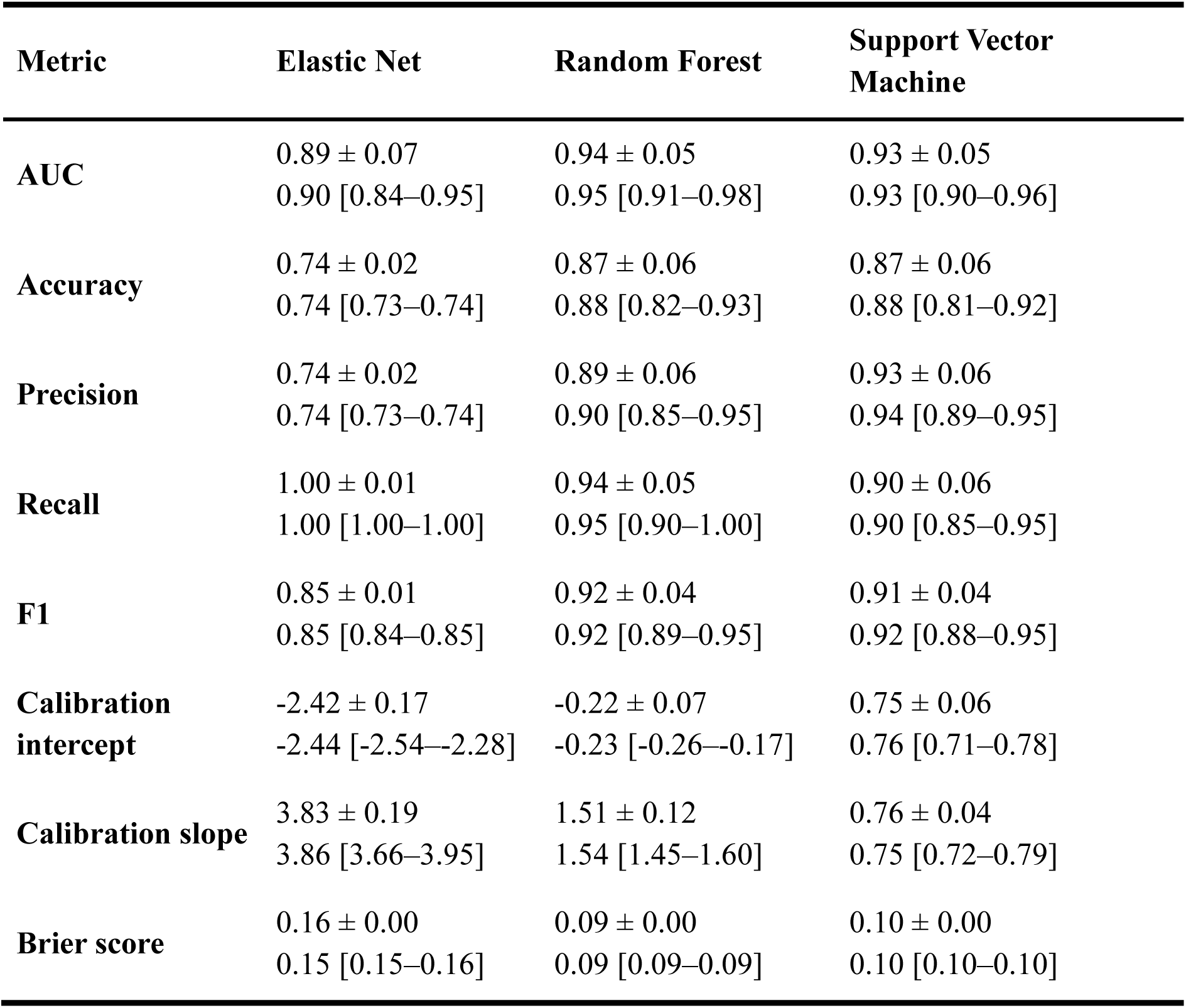
Comparison of predictive performance of the optimally tuned models for predicting rejection: Biopsy whole-transcript expression.

**Supplementary Table 8.**
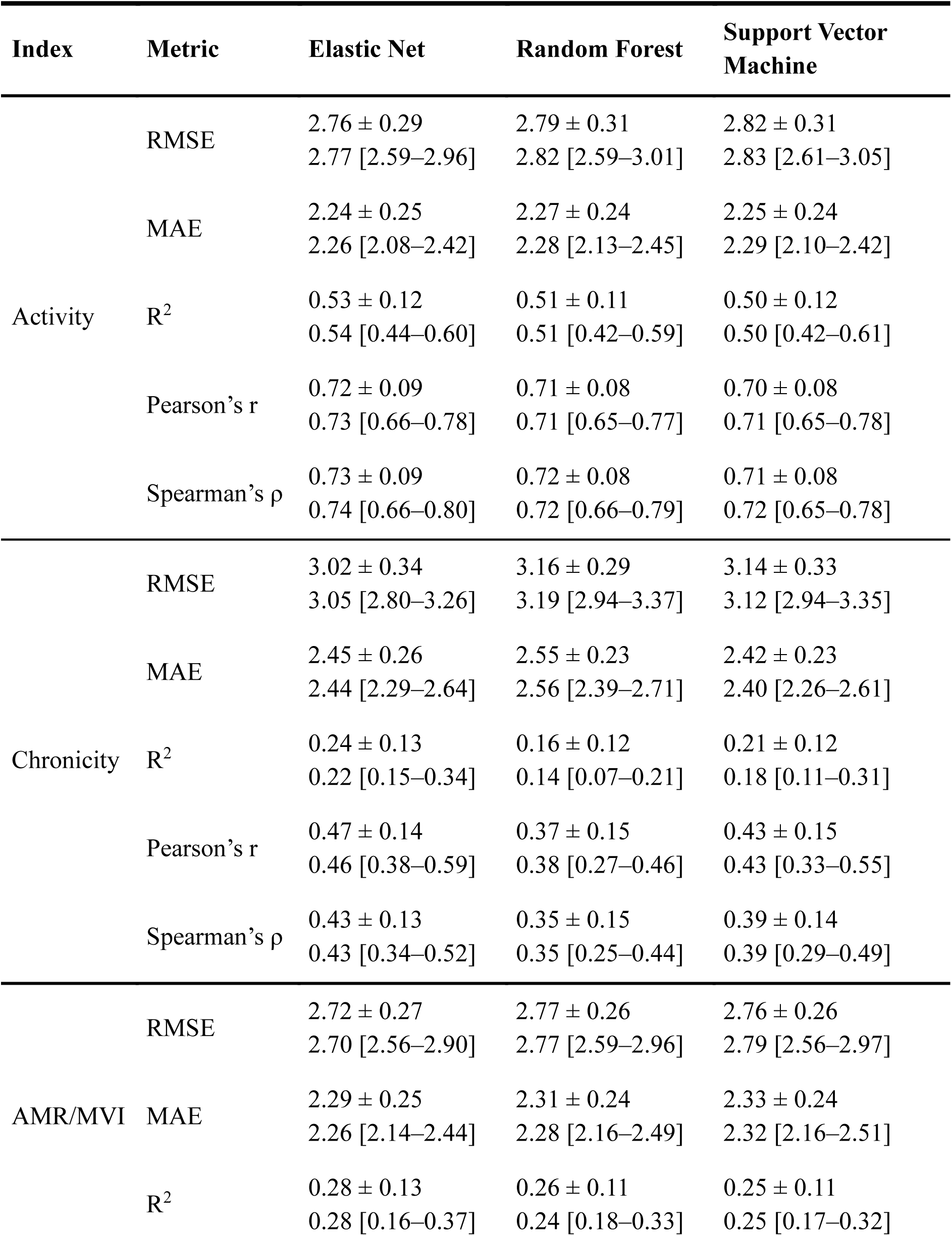
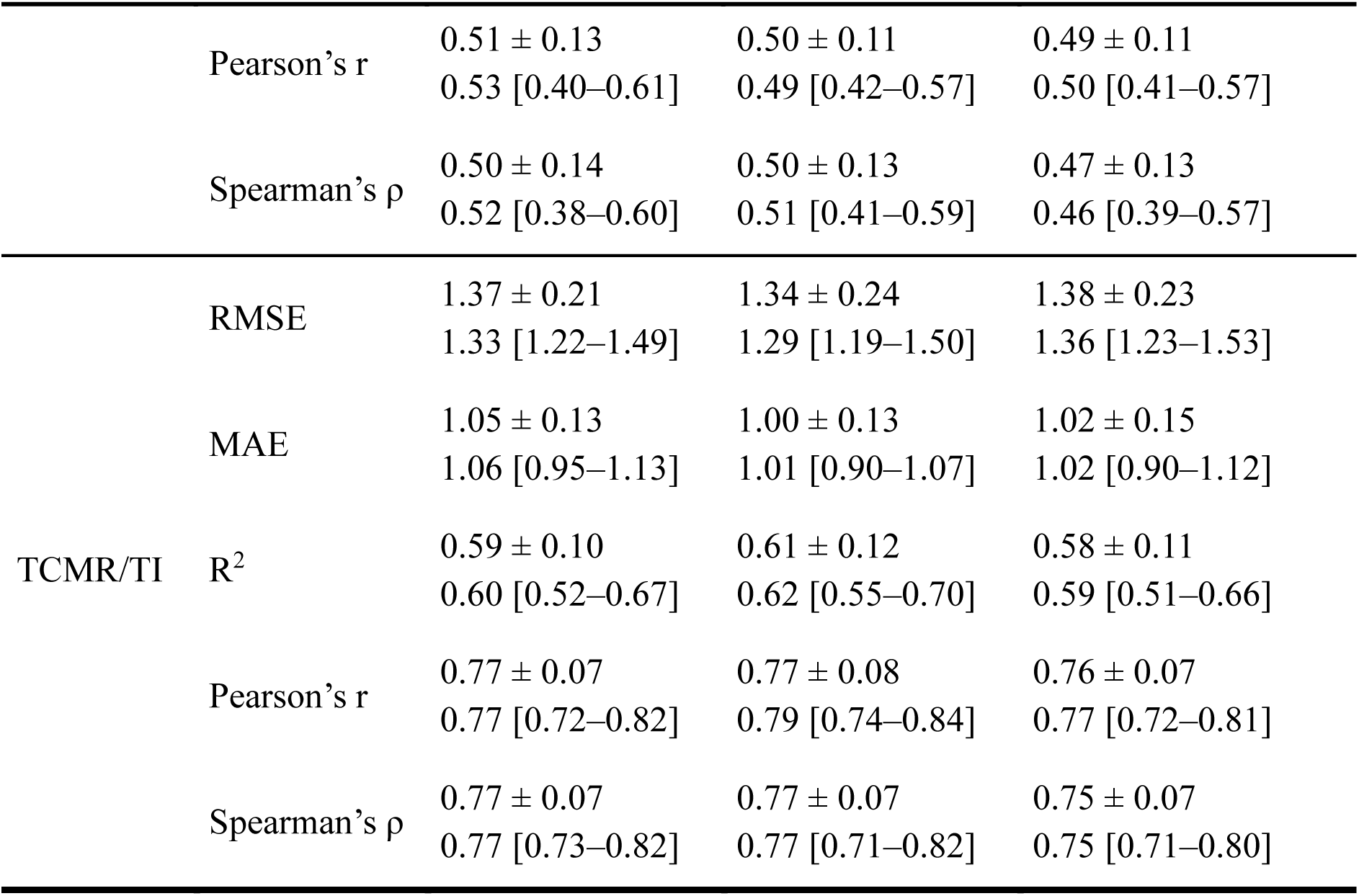
Comparison of predictive performance of the optimally tuned models for predicting continuous index: Biopsy gene-panel expression.

**Supplementary Table 9.**
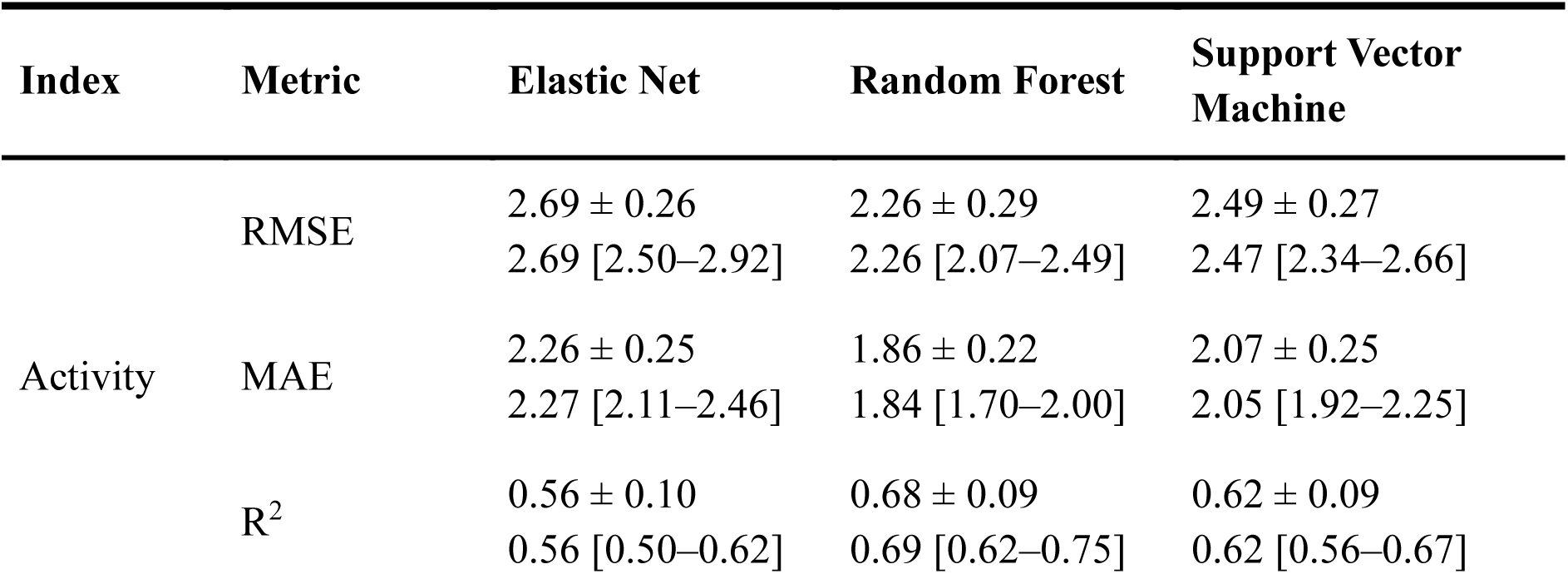
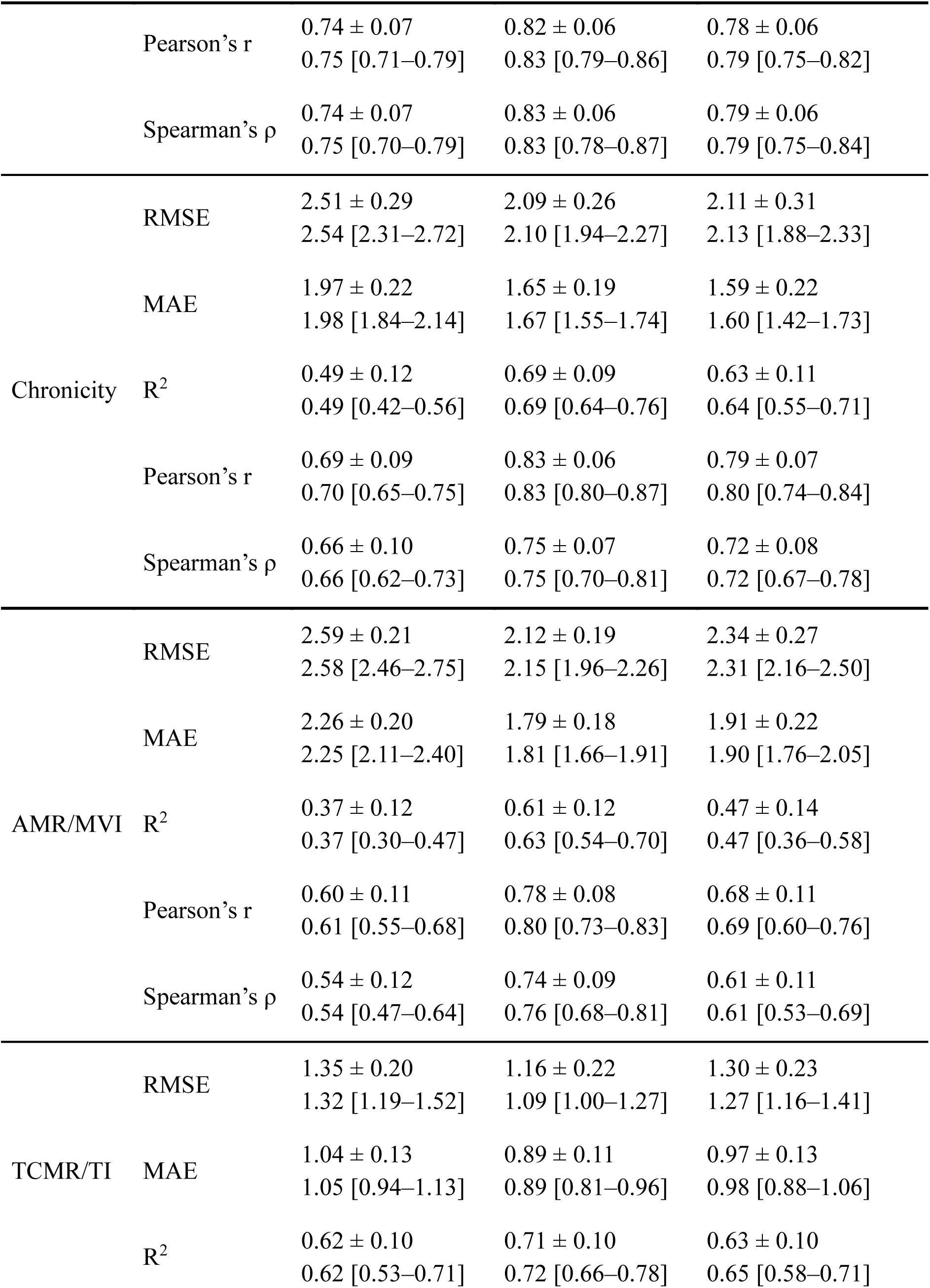
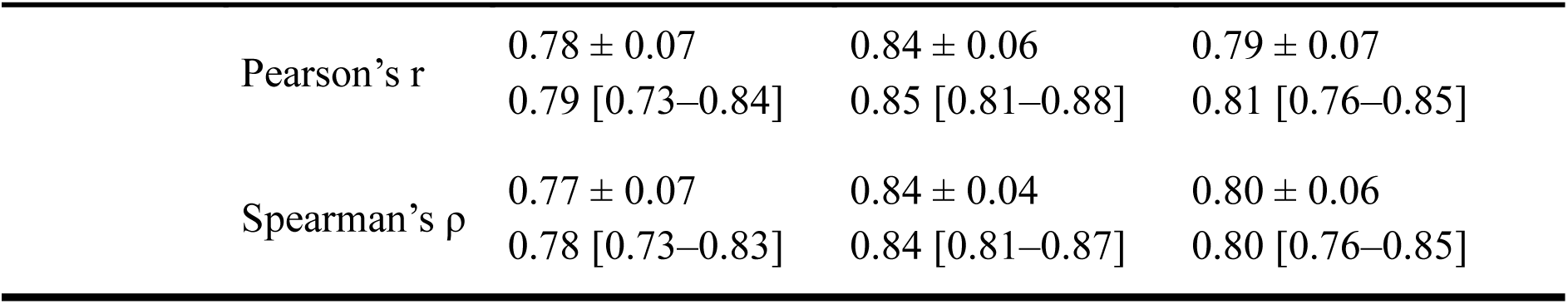
Comparison of predictive performance of the optimally tuned models for predicting continuous index: Biopsy whole-transcript expression.

**Supplementary Figure 1.**
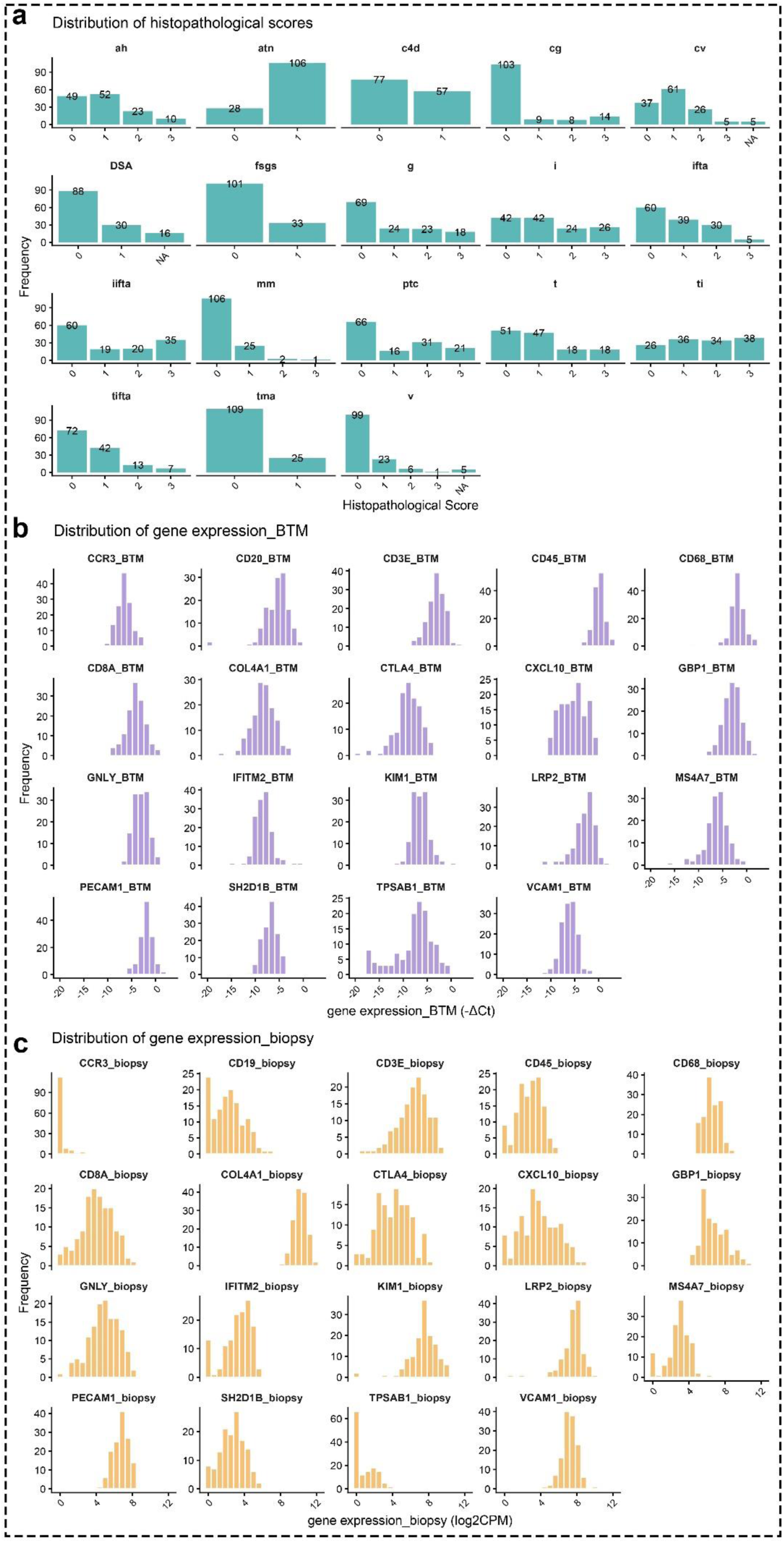
Distribution of histopathological scores and gene expression. (a) Bar plots show the distribution of individual histopathological lesion scores across the study cohort. The x-axis represents the score assigned to each lesion, and the y-axis represents the number of patients with each score. (b) Histograms show the distribution of BTM-derived gene expression. (c) Histograms show the distribution of biopsy-derived gene expression levels.

**Supplementary Figure 2.**
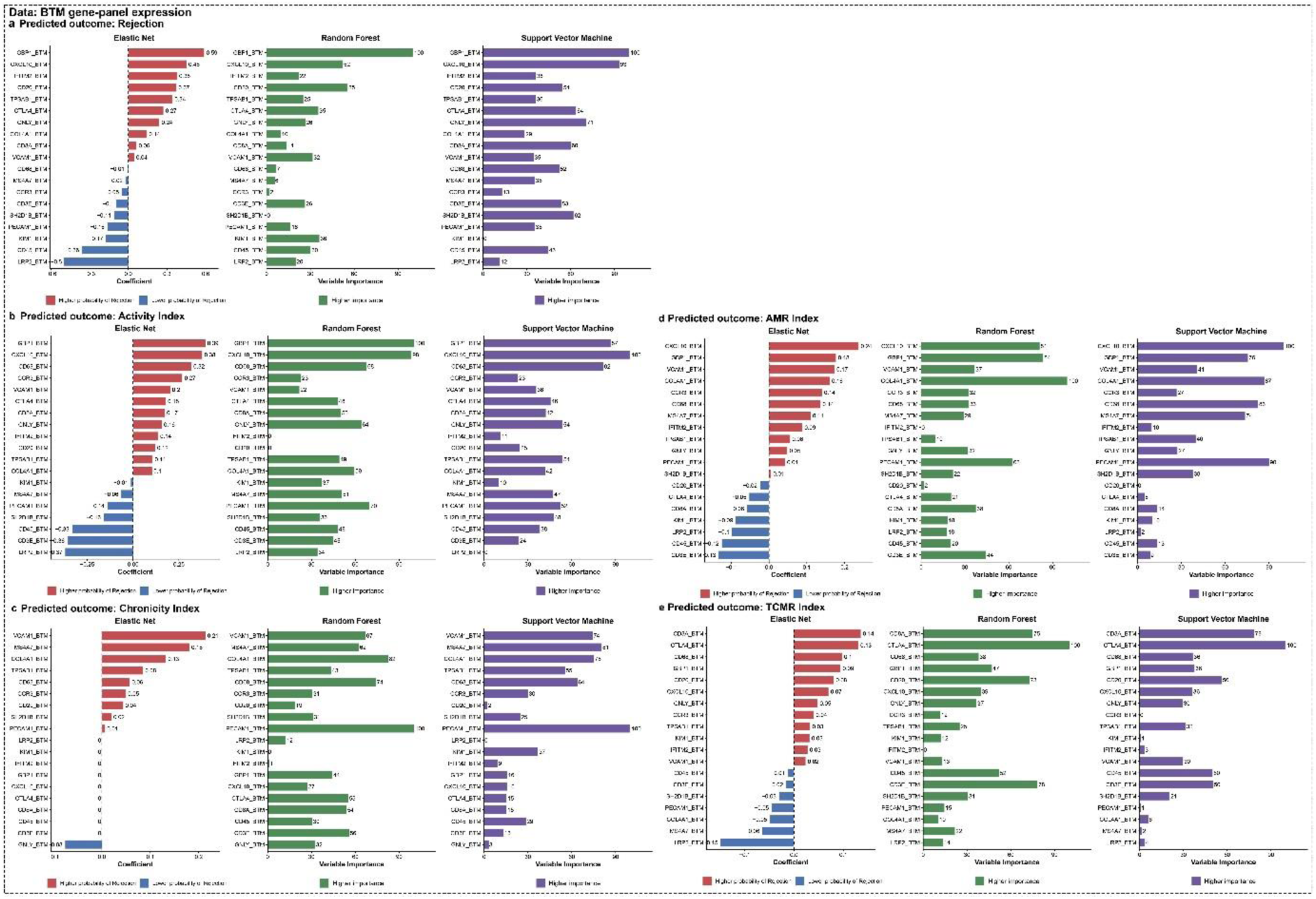
Predictor effects across machine learning models for binary diagnostic classification and continuous index prediction using BTM gene-panel expression. Predictor effects and variable importance are shown for Elastic Net, Random Forest, and Support Vector Machine (SVM) models across (a) rejection, (b) activity index, (c) chronicity index, (d) AMR/MVI index, and (e) TCMR/TI index outcomes. Elastic Net coefficients indicate the direction and magnitude of predictor effects, with positive and negative coefficients representing higher and lower predicted outcomes, respectively. For Random Forest and SVM models, bars represent normalized variable importance.

**Supplementary Figure 3.**
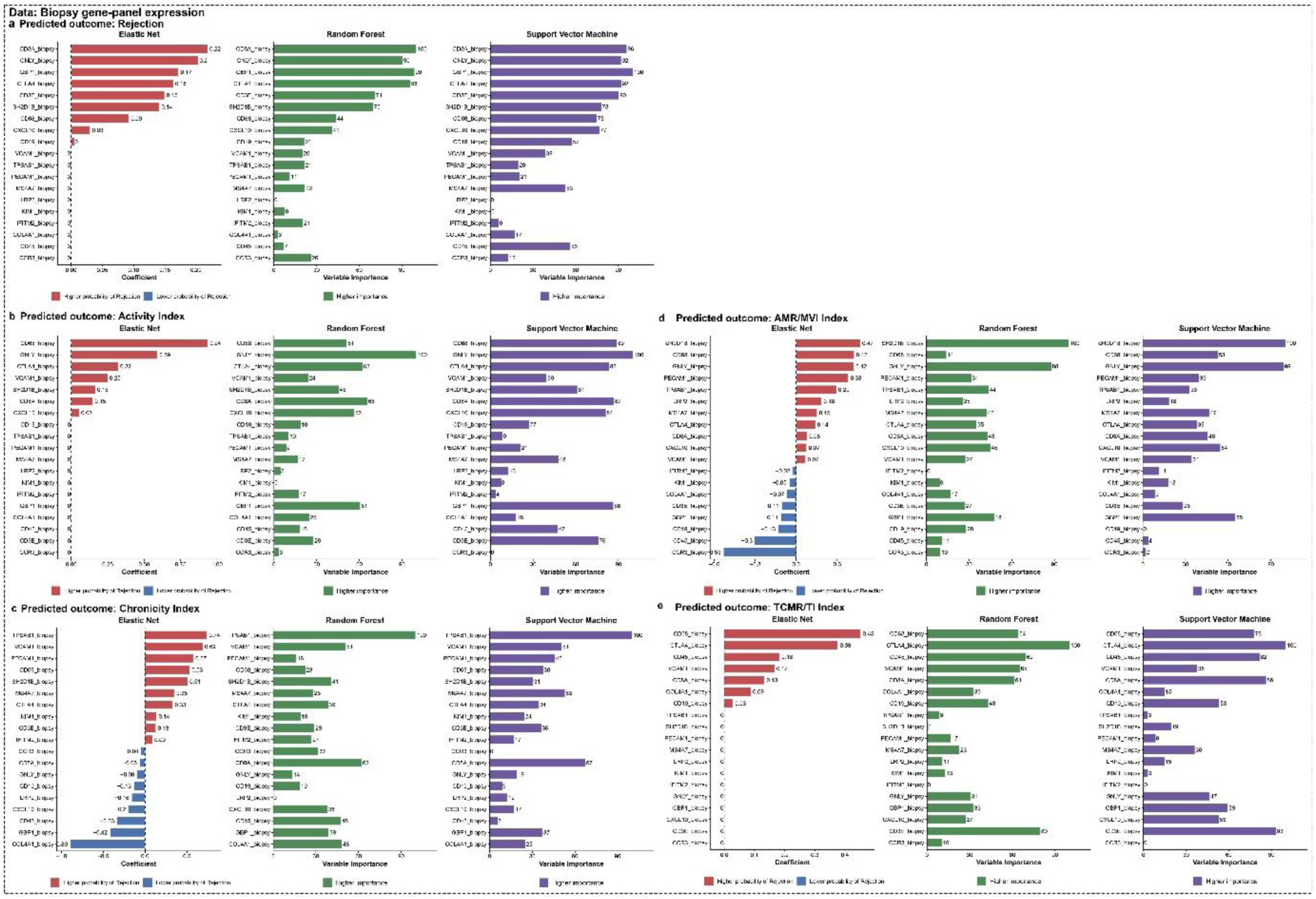
Predictor effects across machine learning models for binary diagnostic classification and continuous index prediction using Biopsy gene-panel expression. Predictor effects and variable importance are shown for Elastic Net, Random Forest, and Support Vector Machine (SVM) models across (a) rejection, (b) activity index, (c) chronicity index, (d) AMR/MVI index, and (e) TCMR/TI index outcomes. Elastic Net coefficients indicate the direction and magnitude of predictor effects, with positive and negative coefficients representing higher and lower predicted outcomes, respectively. For Random Forest and SVM models, bars represent normalized variable importance.

**Supplementary Figure 4.**
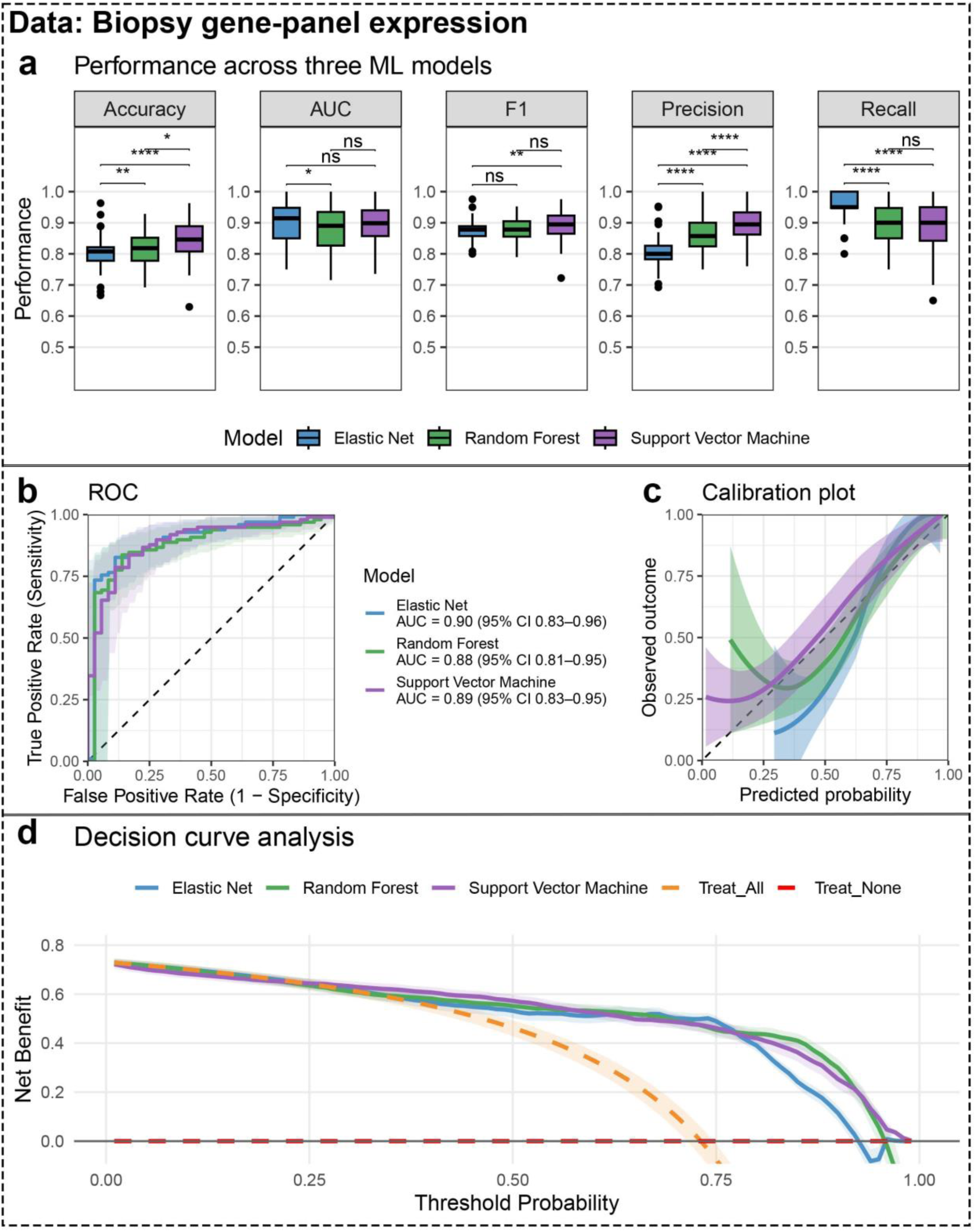
Performance of machine-learning models for binary diagnostic classification using Biopsy gene-panel expression. a, Comparison of classification performance across Elastic Net, Random Forest, and SVM-RBF models, evaluated by accuracy, AUC, F1 score, precision, and recall. Model performance was evaluated using 20 repetitions of five-fold cross-validation. Within each repetition, out-of-fold predictions from the five held-out folds were combined to obtain a single performance estimate, yielding 20 matched estimates per model. Pairwise differences between models were assessed using paired Wilcoxon signed-rank tests across matched repetitions. Statistical significance is indicated above the pairwise comparisons; ns, not significant. **b,** Out-of-fold predicted probabilities were averaged across cross-validation repetitions for each patient, and the resulting patient-level probabilities were used to construct the ROC curves. Shaded regions represent pointwise 95% confidence bands for the ROC curves, estimated using bootstrap resampling. AUCs are reported with 95% confidence intervals calculated using DeLong’s method. **c,** Calibration curves showing the relationship between the averaged patient-level out-of-fold predicted probabilities and observed outcomes for each model. The dashed diagonal line indicates perfect calibration, and shaded areas represent pointwise 95% confidence intervals around the smoothed calibration curves. **d,** Decision curve analysis showing the net benefit of the Elastic Net, Random Forest, and SVM models across a range of threshold probabilities. The dashed orange and red lines indicate the strategies of treating all patients and treating no patients, respectively.

**Supplementary Figure 5.**
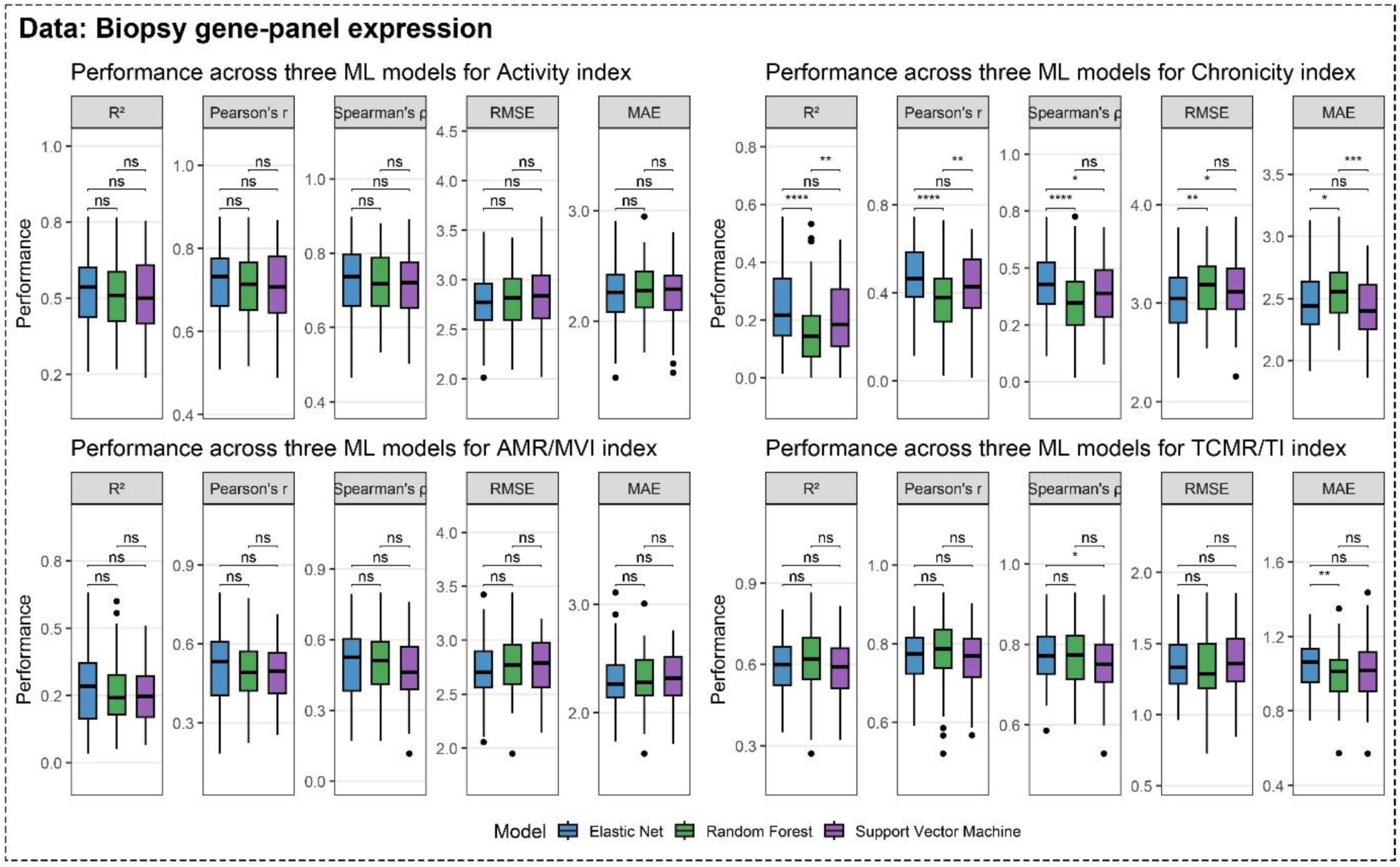
Performance of machine-learning models for continuous index prediction using Biopsy gene-panel expression. Prediction performance of Elastic Net, Random Forest, and SVM-RBF models for the Activity, Chronicity, AMR/MVI, and TCMR/TI indices. Model performance was evaluated using 20 repetitions of five-fold cross-validation. Within each repetition, out-of-fold predictions from the five held-out folds were combined to obtain a single performance estimate, yielding 20 matched estimates per model. Predictive performance was assessed using the coefficient of determination (R²), Pearson’s correlation coefficient (*r*), Spearman’s rank correlation coefficient (ρ), root mean squared error (RMSE), and mean absolute error (MAE). Boxplots show the distributions of performance estimates across the 20 cross-validation repetitions. Pairwise differences between models were assessed across matched repetitions using paired Wilcoxon signed-rank tests. Statistical significance is indicated above the pairwise comparisons; ns, not significant.

**Supplementary Figure 6.**
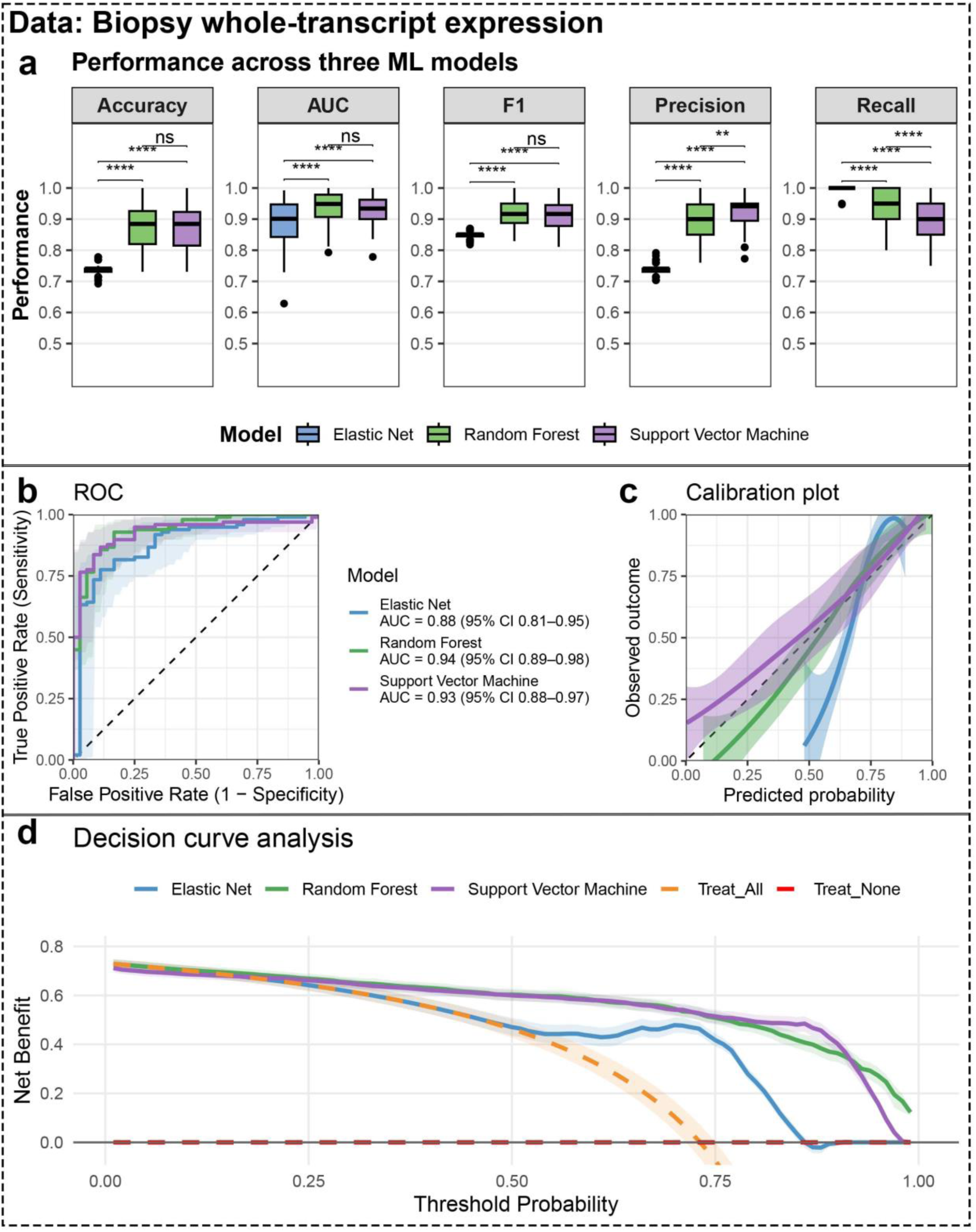
Performance of machine-learning models for binary diagnostic classification using Biopsy whole-transcript expression. a, Comparison of classification performance across Elastic Net, Random Forest, and SVM-RBF models, evaluated by accuracy, AUC, F1 score, precision, and recall. Model performance was evaluated using 20 repetitions of five-fold cross-validation. Within each repetition, out-of-fold predictions from the five held-out folds were combined to obtain a single performance estimate, yielding 20 matched estimates per model. Pairwise differences between models were assessed using paired Wilcoxon signed-rank tests across matched repetitions. Statistical significance is indicated above the pairwise comparisons; ns, not significant. **b,** Out-of-fold predicted probabilities were averaged across cross-validation repetitions for each patient, and the resulting patient-level probabilities were used to construct the ROC curves. Shaded regions represent pointwise 95% confidence bands for the ROC curves, estimated using bootstrap resampling. AUCs are reported with 95% confidence intervals calculated using DeLong’s method. **c,** Calibration curves showing the relationship between the averaged patient-level out-of-fold predicted probabilities and observed outcomes for each model. The dashed diagonal line indicates perfect calibration, and shaded areas represent pointwise 95% confidence intervals around the smoothed calibration curves. **d,** Decision curve analysis showing the net benefit of the Elastic Net, Random Forest, and SVM models across a range of threshold probabilities. The dashed orange and red lines indicate the strategies of treating all patients and treating no patients, respectively.

**Supplementary Figure 7.**
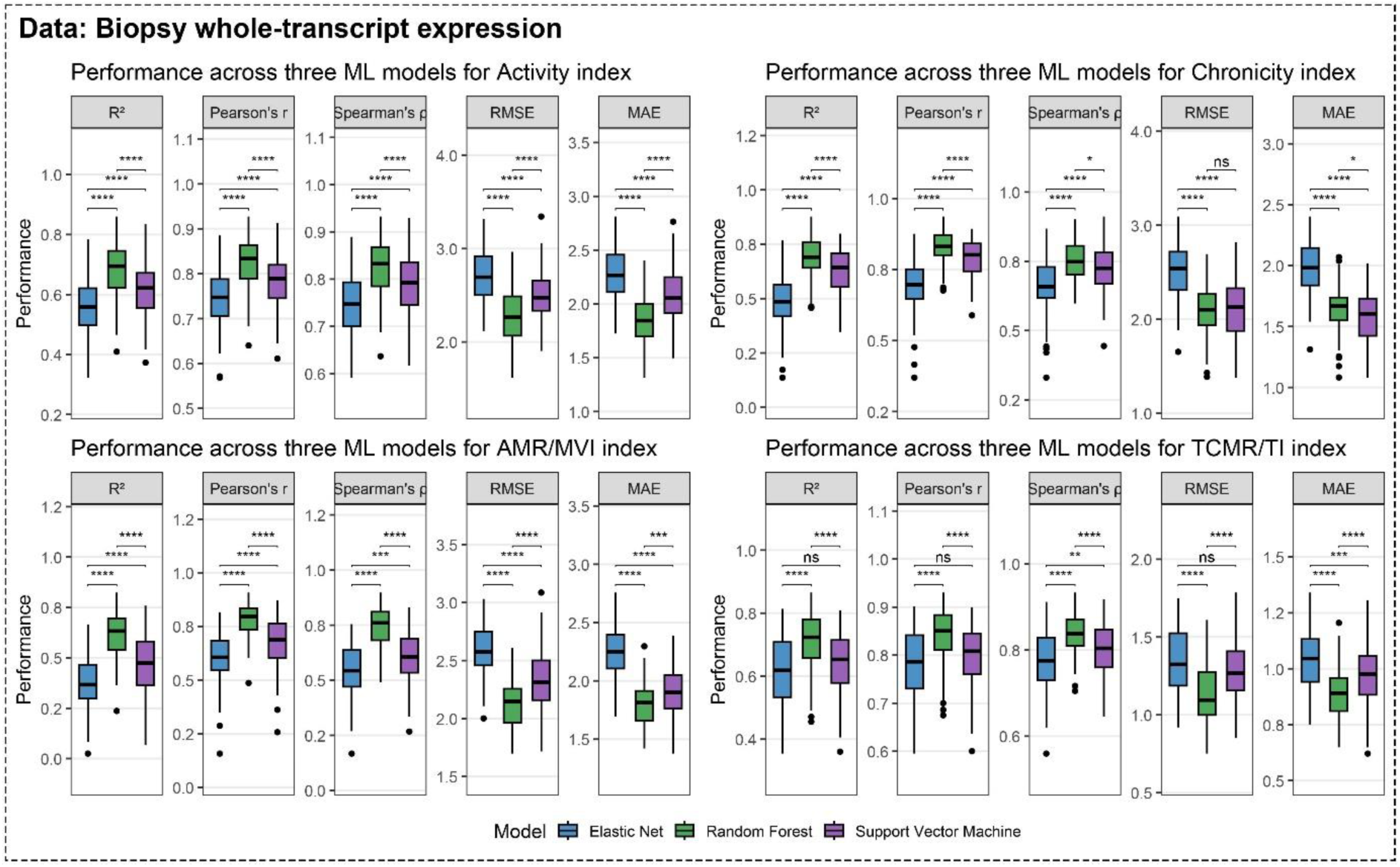
Performance of machine-learning models for continuous index prediction using Biopsy whole-transcript expression. Prediction performance of Elastic Net, Random Forest, and SVM-RBF models for the Activity, Chronicity, AMR/MVI, and TCMR/TI indices. Model performance was evaluated using 20 repetitions of five-fold cross-validation. Within each repetition, out-of-fold predictions from the five held-out folds were combined to obtain a single performance estimate, yielding 20 matched estimates per model. Predictive performance was assessed using the coefficient of determination (R²), Pearson’s correlation coefficient (*r*), Spearman’s rank correlation coefficient (ρ), root mean squared error (RMSE), and mean absolute error (MAE). Boxplots show the distributions of performance estimates across the 20 cross-validation repetitions. Pairwise differences between models were assessed across matched repetitions using paired Wilcoxon signed-rank tests. Statistical significance is indicated above the pairwise comparisons; ns, not significant.

## Supplementary Methods

### Predictive modeling framework

#### Definition of modeling datasets and reference models

Three modeling datasets were used. The primary BTM gene-panel models were developed using qPCR measurements of the predefined gene panel in BTM samples. As molecular references, biopsy gene-panel models were developed using the corresponding BTM genes extracted from the biopsy TempO-Seq data, whereas biopsy whole-transcript models were developed using all transcripts measured by TempO-Seq in the biopsy samples. The biopsy-derived models were included as molecular reference models to assess the predictive capacity of the predefined gene panel in biopsy tissue and to determine whether inclusion of additional transcriptomic features could potentially improve predictive performance in the future.

#### Development of prediction models

The entire cohort was used to develop prediction models. Model development and internal validation were performed using repeated five-fold cross-validation with 20 repetitions, resulting in 100 resampling iterations. Prediction models were developed using three supervised machine learning algorithms: Elastic Net (EN), Random Forest (RF), and Support Vector Machine with a radial basis function kernel (SVM-RBF). All models were trained using the same resampling strategy defined in the caret training control object to ensure fair comparison across algorithms.

Hyperparameters were optimized using predefined grid searches. For EN, the mixing parameter α was evaluated from 0 to 1 in increments of 0.1, where α = 0 corresponds to ridge regression and α = 1 to lasso regression. The regularization parameter λ was evaluated across 50 logarithmically spaced values ranging from 10^−3^ to 10^2^. Predictors were centered and scaled before model fitting. For RF, the number of predictor variables randomly considered at each split (mtry) was evaluated at values of 2, 4, 6, 8, 10, 12, 14, 16, and 18. Each model consisted of 500 trees, and variable importance was calculated. For SVM-RBF, predictors were centered and scaled before model fitting. The radial basis function kernel parameter σ was evaluated across 10 logarithmically spaced values ranging from 10^−3^ to 10^0^, while the cost parameter C was evaluated at 0.01, 0.1, 1, 10, and 100.

For analyses using whole-transcriptome expression data, feature filtering and selection were performed using the training set only to avoid information leakage. Genes with near-zero variance across the training samples were first removed using the nearZeroVar function in the caret package. Genes with zero variance after this filtering step were additionally excluded.

All subsequent feature-selection procedures were performed on the remaining genes. Recursive feature elimination (RFE) was then performed separately with RF and SVM-RBF as the underlying classifiers to select the optimal feature subset for each model. Candidate feature-set sizes of 19, 60, 125, 250, and 500 genes were evaluated. For RF-based RFE, RF models containing 1,000 trees were used to rank and select predictors. For SVM-RBF-based RFE, a linear SVM-RBF was used for feature ranking and selection. The feature subset yielding the highest cross-validated area under the receiver operating characteristic curve (ROC AUC) was retained separately for each modeling algorithm.

#### Evaluation of model performance

Model performance was evaluated using the out-of-fold predictions generated from the optimal hyperparameter configuration (best-tuned model) identified during repeated 5-fold cross-validation. For the binary diagnosis, model discrimination was assessed using AUC, accuracy, precision, recall, and F1-score, with repeated out-of-fold predictions averaged for each observation to generate ROC curves. Model calibration was assessed using calibration plots and the calibration intercept and slope, which evaluate systematic bias in predicted risks and the spread of predictions, respectively. Overall probabilistic prediction accuracy was assessed using the Brier score. The clinical utility of each binary prediction model was evaluated using decision curve analysis (DCA) across a range of clinically relevant probability thresholds^18^. Net benefit was compared with two default approaches in patients undergoing biopsy for suspected rejection: assuming that all biopsies represent rejection versus assuming that none represent rejection. For the continuous indices, predictive performance was assessed using the coefficient of determination (R²), root mean square error (RMSE), mean absolute error (MAE), with Pearson (r) and Spearman (ρ) correlation coefficients additionally used to assess the association between predicted and observed values.

#### Interpretation of model predictions

Gene-level coefficients or variable importance scores were extracted from the final models fitted with the optimal hyperparameters. For the EN model, coefficients were extracted and odds ratios were calculated by exponentiating the coefficient estimates. For the RF and SVM-RBF models, scaled variable importance scores were obtained using the variable importance function in the caret package. Coefficients and importance scores were visualized to identify the genes contributing most strongly to each model.

